# Investigating the transparency of reporting in two-sample summary data Mendelian randomization studies

**DOI:** 10.1101/2021.10.15.21264972

**Authors:** Benjamin Woolf, Nina Di Cara, Christopher Moreno-Stokoe, Veronika Skrivankova, Katie Drax, Julian P.T. Higgins, Gibran Hemani, Marcus R. Munafò, George Davey Smith, James Yarmolinsky, Rebecca C. Richmond

## Abstract

**Background:** Two-sample Mendelian randomization (2SMR) is an increasingly popular epidemiological method that uses genetic variants as instruments for making causal inferences. Clear reporting of methods employed in such studies is important for evaluating their underlying quality. However, the quality of methodological reporting of 2SMR studies is currently unclear.

**Objectives:** We aimed to assess the reporting quality of studies that used MR-Base, one of the most popular platforms for implementing 2SMR analysis.

**Methods:** We created a bespoke reporting checklist to evaluate reporting quality of 2SMR studies. We then searched Web of Science Core Collection, PsycInfo, MEDLINE, EMBASE and Google Scholar citations of the MR-Base descriptor paper to identify published MR studies that used MR-Base for any component of the MR analysis. Study screening and data extraction were performed by at least two independent reviewers.

**Results:** 87 studies were included in the primary analysis. Reporting quality was generally poor across studies with a mean of 53% (SD = 14%) of items reported in each study. Many items required for evaluating the validity of key assumptions made in MR were poorly reported: only 44% of studies provided sufficient details for assessing if the genetic variant associates with the exposure (‘relevance’ assumption), 31% for assessing if there are any variant-outcome confounders (‘independence’ assumption), 89% for the assessing if the variant causes the outcome independently of the exposure (‘exclusion restriction’ assumption), and 32% for assumptions of falsification tests. We did not find evidence of a change in reporting quality over time or a difference in reporting quality between studies that used MR-Base and a random sample of MR studies that did not use this platform.

**Discussion:** The quality of reporting of two-sample Mendelian randomization studies in our sample was generally poor. Journals and researchers should implement the STROBE-MR guidelines to improve reporting quality.

**Other: Funding:** ESRC, CRUK, MRC, John Climax Benevolent Fund, University of Bristol, and the Wellcome Trust. Registration: This study pre-registered on the OSF, and the protocol can be found at DOI 10.17605/OSF.IO/NFM27

## Introduction

Mendelian randomization (MR) is an epidemiological approach to causal inference which uses germline genetic variants strongly associated with exposures of interest to appraise the effect of those exposures on one or more outcomes [1]. In a traditional “one-sample” MR design, information on exposure, outcome, and genetic instruments are collected in a single sample, and the effect of the exposure on the outcome estimated using only these data. Two-sample Mendelian randomization (2SMR) instead uses two studies with information on the association between the genetic instrument(s) and the exposure (Sample 1) and the genetic instrument(s) and the outcome (Sample 2), under the assumption that both samples are representative of the same underlying population, to calculate an effect estimate [2]. Because 2SMR only requires information on each genotype-phenotype association, summary statistics from two genome-wide association studies (GWASs) often provide sufficient information to implement the analysis. The increase in publicly accessible summary statistics from GWAS has vastly facilitated the application of 2SMR. Since the summary statistics generally come from previously published data, there is no requirement to apply for and access individual-level data or to perform data cleaning. This in turn makes performing MR analysis more rapid. The emergence of platforms automating the curation of GWAS summary statistics, and statistical packages for performing 2SMR (e.g. MR-Base/TwoSampleMR [3], SMR [4]) and MendelianRandomization [5]) has also facilitated the growth in studies employing this method by reducing the complexity of implementing 2SMR analysis [6].

IEU OpenGWAS (https://gwas.mrcieu.ac.uk/) is a curated, open source, GWAS summary statistic repository managed by the University of Bristol, with an accompanying R package (TwoSampleMR) and web platform (MR-Base) for performing 2SMR analysis[3]. At the start of 2021, the repository contained nearly 40,000 GWASs across many categories of traits including biomarkers, clinical conditions, and behavioural traits. The R package and web platform, hereafter collectively referred to as MR-Base, can be used to perform many stages of a 2SMR analysis, including harmonization of data across datasets and commonly used sensitivity analyses, using data available from a linked data repository or uploaded by the user. The integration of large GWAS repositories and an easy-to-implement analysis package makes MR-Base one of the most popular tools for performing two-sample MR analysis. For example, at the time of writing, in June 2021, the descriptor paper for MR-Base[3], and associated R-package, had between 2 and 3 times more citations in Google Scholar than the citations for an alternative package’s description paper published at a similar time[7], and for the 12 months prior to 06/2021 MR-Base received around 95 million API (Application Programming Interface) requests[8].

While MR-Base increases the breadth and speed at which 2SMR analyses can be performed, it might also lead to discrepancies in their design, conduct and reporting. The accessibility of such resources may permit analyses to be performed without careful consideration of the analytical choices that are being made or the assumptions inherent in the approach. Further, the abundance of genetic instruments, datasets and methods available for use might influence the robustness of study results [6,9]. For example, easily accessible GWAS data of uncertain quality may be used in an analysis without the ability to examine or correct for bias in the underlying dataset. Even if the methodological design of the underlying GWAS used in a 2SMR analysis is robust, the ability to rapidly extract summary statistics directly from a data repository, rather than from the original GWAS itself, may encourage insufficient assessment and reporting of the methods used in the GWAS (e.g. with regard to selection or phenotyping procedures).These factors may encourage the generation of spurious and/or non-reproducible results which could present a possible threat to the robustness and reliability of the literature. Automation of MR analysis permits rapid assessment of numerous potentially causal relationships which may also encourage “data fishing” (e.g. examining various hypotheses and reporting only “positive findings”) or selectively “cherry-picking” results from sensitivity analyses.

Given concern that such platforms may facilitate poor quality or poorly reported research [10], there is therefore a need to systematically appraise the quality of reporting in 2SMR studies in order to make an assessment of the potential quality and rigour of the analysis performed. Assessing the transparency of reported studies is also a necessary part of assessing their risk of bias and is a requirement for good attempts at replication. Unless the methods and assumptions of a study are explicitly stated, there will likely be a large number of “researcher degrees of freedom” leading to variability and inconsistency in findings [11]. A final benefit of systematically assessing transparency is that doing so requires the creation of an explicit checklist for the appraisal of the reporting quality in MR studies.

This project aimed to appraise the transparency in reporting of two-sample Mendelian randomization analyses. We therefore examined the reporting quality of 2SMR analyses that have been performed using the MR-Base statistical package (TwoSampleMR in R) and data repository using a checklist developed specifically for this study.

## Methods

### Development of the reporting quality checklist

We developed a bespoke checklist to examine the quality of reporting of 2SMR studies because none of the existing MR-reporting reviews or guidelines [1, 12-15] were specifically tailored to 2SMR, and therefore did not include items on 2SMR specific assumptions and requirements, such as data harmonisation. To inform the checklist, BW reviewed papers describing assumptions and sources of bias in MR analyses in addition to papers presenting reporting checklists for MR and/or instrumental variable studies. BW produced a first draft of the checklist which was then amended iteratively based on feedback from JY and RR and piloted on three 2SMR studies. The development of the checklist was also informed by discussions about the STROBE-MR checklist, which was being developed at the same time by many of this study’s authors.

### Eligibility criteria

The study aimed to assess the quality of reporting of 2SMR studies published in peer-reviewed academic journals. Therefore, any published study that conducted MR and used the MR-Base R package (TwoSampleMR) or online platform (http://www.mrbase.org/) during any component of the MR analysis was eligible for inclusion. Studies were included irrespective of the type of study participants, setting, exposure(s) or outcome(s) being investigated.

### Identification and selection of 2SMR studies

We searched Web of Science Core Collection, PsycInfo, MEDLINE and EMBASE from January 2016, because MR-Base was developed in 2016. The last search date was April 2019 and no language or other constraints were applied. The search terms are provided in the Supplementary Methods.

Studies were also identified by performing a citation search using Google Scholar for citations of the R package: the eLife article [3], the correctly cited bioRxiv preprint[16], incorrectly cited bioRxiv preprint version 1 [17], incorrectly cited bioRxiv preprint version 2 [18], the LD-hub and MR-Base presentation at the 2016 Annual Meeting of the Behaviour-Genetics-Association [19], and references of “www.mrbase.org” in Google Scholar.

Citations retrieved by the search were uploaded onto Rayyan (https://rayyan.qcri.org)[20], a website specifically designed for paper screening in systematic reviews. Rayyan automatically identifies duplicates of citation/abstracts, which were then manually checked for errors. Two reviewers (BW, ND) screened the abstracts and titles for relevance using the eligibility criteria. Studies identified as potentially relevant had their full text screened. When both reviewers agreed that a study met the eligibility criteria it was included in the review. Initial disagreements were discussed between ND and BW, with unresolved items arbitrated by a third researcher (JY or RR).

### Data collection process

We developed a standardised data extraction form in advance of data collection. The form required information on each paper’s author(s), date of publication, title, and quality of reporting across all items included in the reporting quality checklist. Each item in the checklist was graded as having been reported (1), not having been reported (0), or not being applicable to the study (NA), based on the reviewer’s opinion of the detail of reporting. Information to support each of these assessments was also collected (e.g. as quotations from the papers). The form was pilot tested on three 2SMR papers that did not use MR-Base to ensure that the same studies would not be included in the final review.

Data extraction and grading for each study were performed by two independent reviewers (from among five reviewers: BW, ND, CMS, JY, and RR) to minimise transcription errors. The data extraction forms were then combined and checked for errors in extraction and disagreements. Disagreements were arbitrated by the reviewers re-reading the paper and coming to a joint conclusion.

To check that unique studies were only included once, studies that shared at least one author were compared based on similarity of study population, date, and methodology. Duplicate studies were treated as a single study in the analysis. Because this review aimed to assess the quality of reporting of 2SMR studies in the published literature, no attempt was made to contact study authors for further information.

### Analysis

Defining a “study” as an individual publication, we calculated a) the percentage of studies reporting each individual item in the checklist; b) the percentages of studies reporting at least 25%, 33%, 50%, 67%, 75% and 100% of all the items in the checklist; and c) the overall mean percentage of items reported across all studies. Percentages excluded studies rated as not applicable for any specific item.

Since studies analysing multiple phenotypes (e.g. phenome-wide scans) may not report all exposure-outcome associations with equal detail, we also evaluated studies based on whether they were a “multi-phenotype study” or not, defining “multi-phenotype” as a study with ≥ 1 exposure and ≥ 10 outcomes or a study with ≥ 10 exposures and ≥ 1 outcome. For brevity, “multi-phenotype” studies are referred to as “≥10 phenotype” studies, non-multi-phenotype studies are referred to as “<10 phenotype studies”. For multi-phenotype studies, the items in the GWAS section of the checklist were calculated as the mean reporting quality across all included GWASs used in the MR analysis.

We undertook four supplementary analyses:

1. We compared changes in the level of reporting (mean percentage of all items reported) across studies by the year of publication.
2. BW and GH classified each item as having information that would be available to all users (i.e. the information required to report the item is provided by the output of both the R package and web platform), information that would be available to some users (i.e. the information required to report the item is available to users of the R package or the web platform, but not both), information that is not given by MR-Base (i.e. the information required to report the item is not available to users of either the R package or the web platform), or as not applicable to the MR-Base analysis (e.g., for items defining the study and/or articulating the question) (Table 1). We then compared the level of reporting according this classification.

**Table 1:**
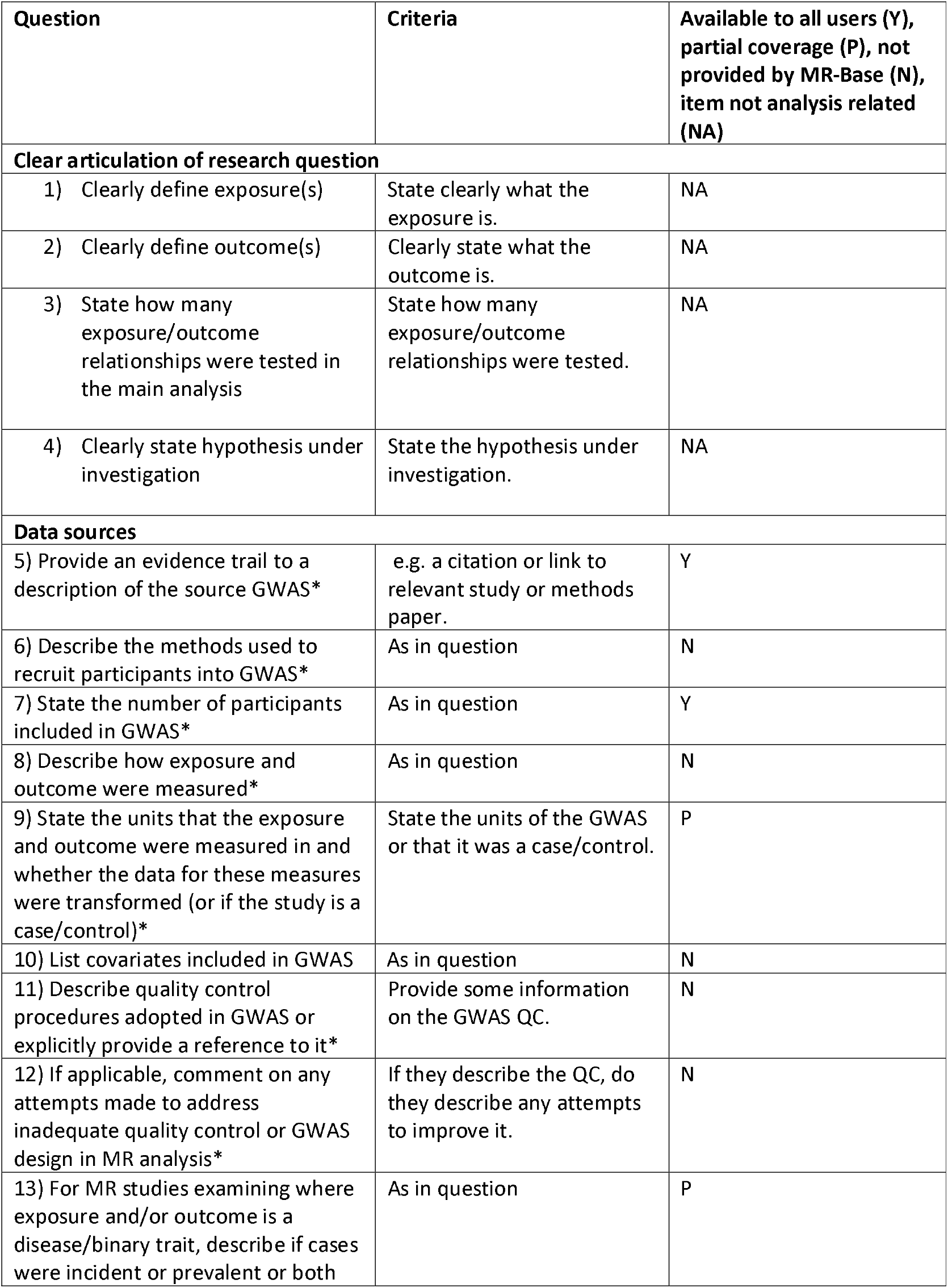
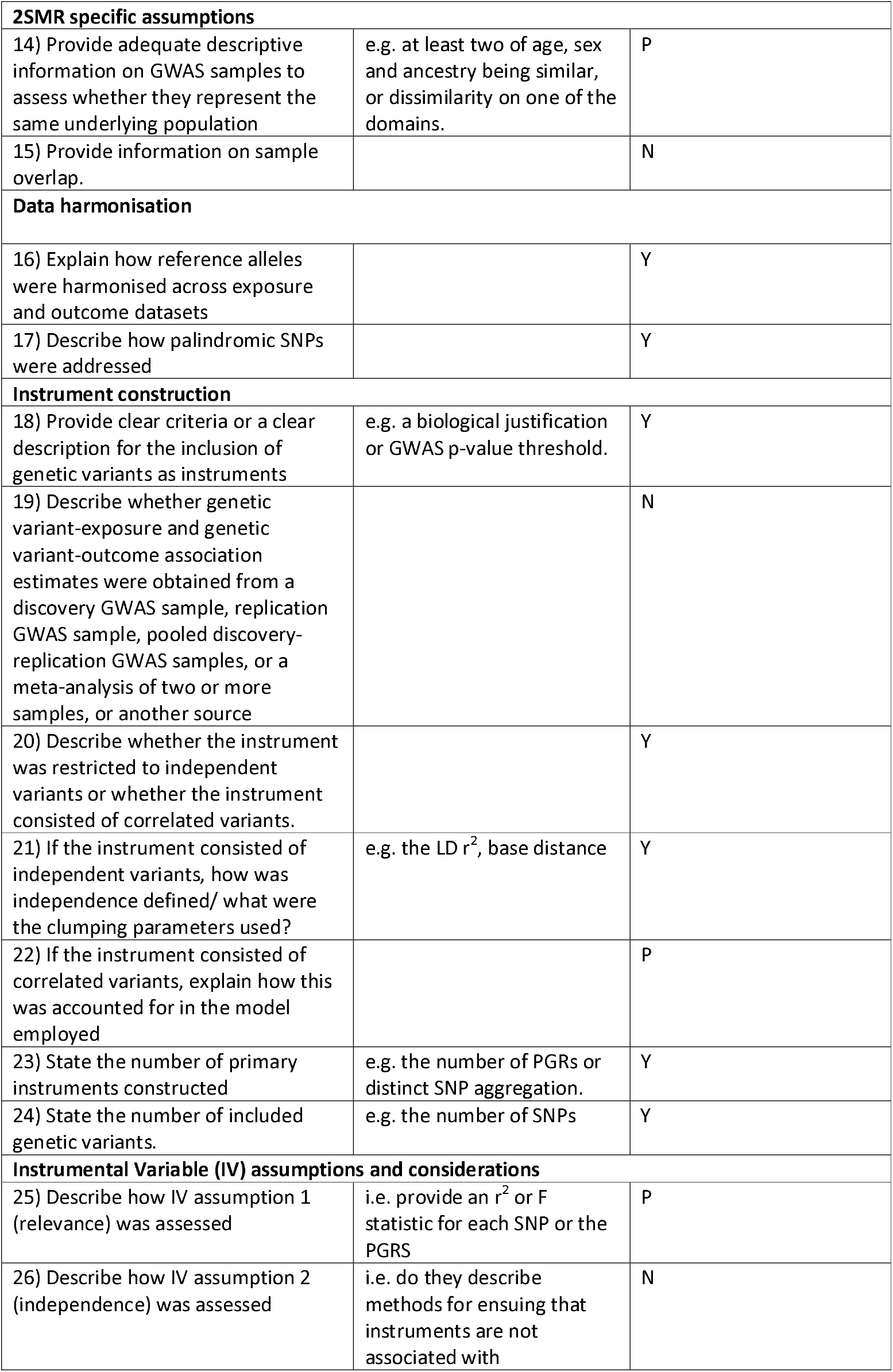
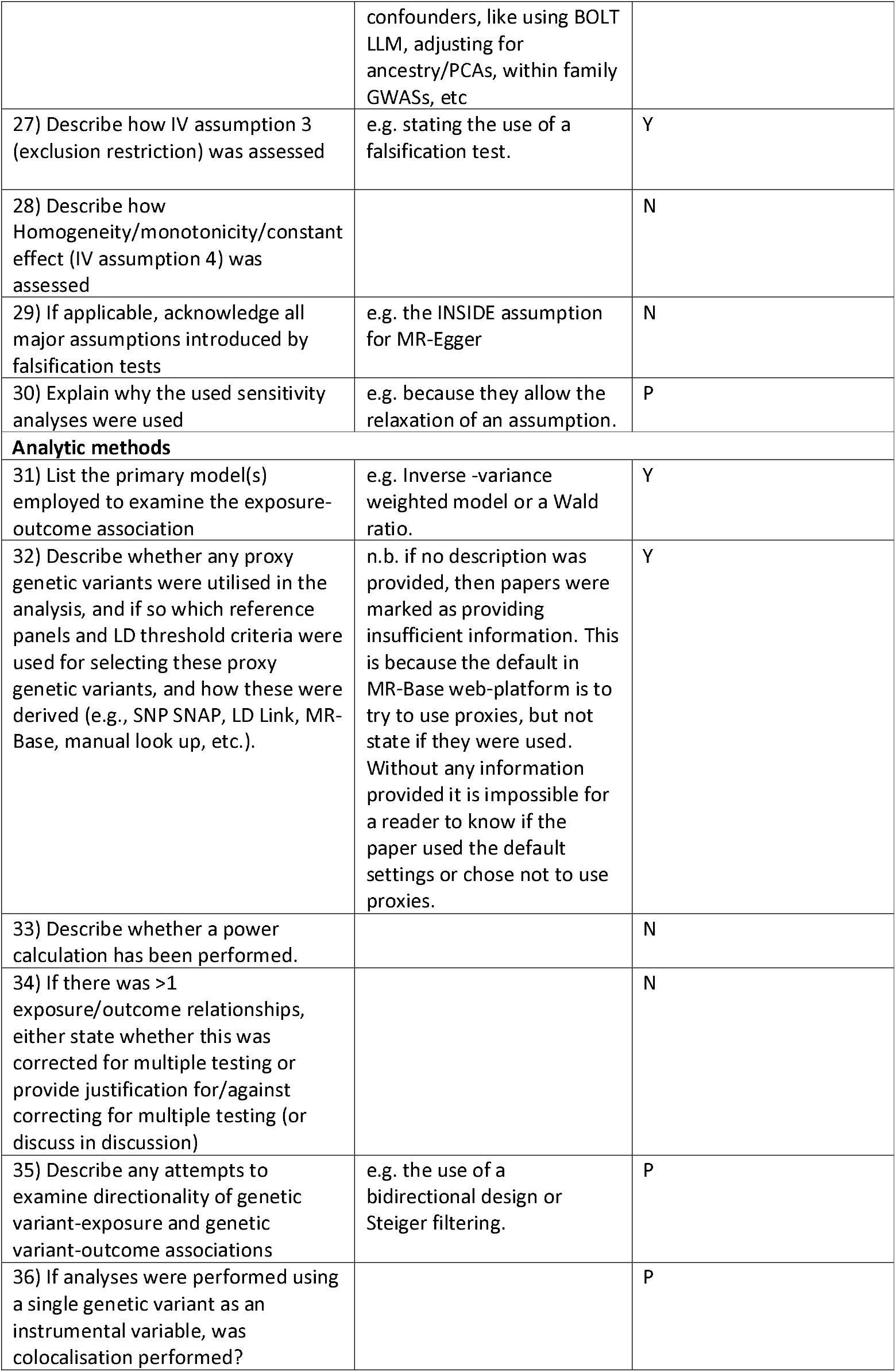
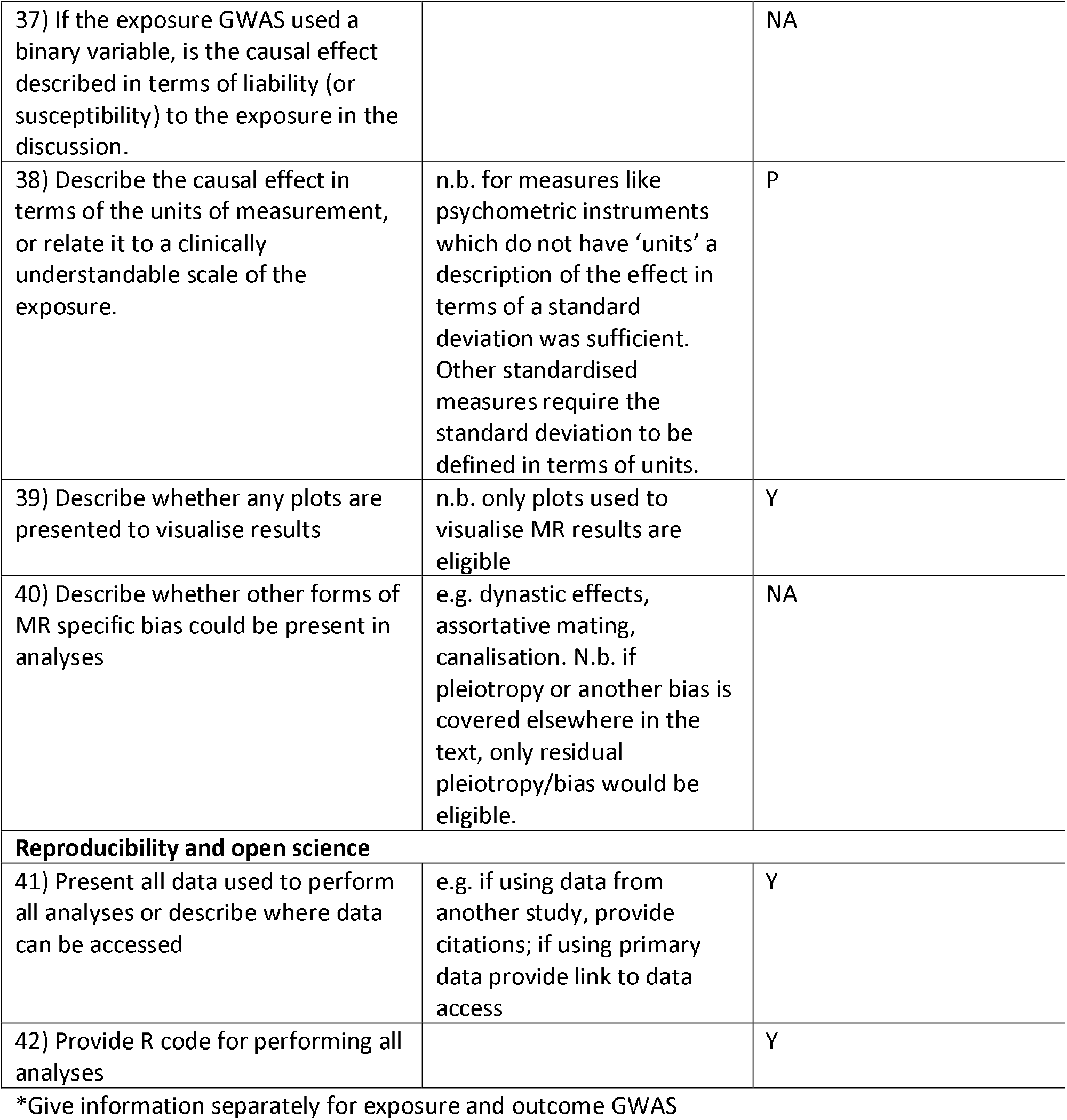
Reporting checklist
3. We compared the level of reporting according to method of citation, classified as explicitly citing the R-package or not citing the R package.
4. While drafting the STROBE-MR extension [21], VS, RR, BW, and JY examined a random sample of MR papers using the original STROBE checklist: A description of the methods used in this review can be found in the supplementary methods. To explore the generalisability of our finds to non-MR-Base papers, we compared the level of reporting between MR-Base and non-MR Base papers for those items that could be harmonised across the reviews. Because this review is a random sample of MR-studies, 95% CIs were calculated to quantify uncertainty due to random sampling error for these estimates.

### Registration

This study, including the first draft of the checklist, was pre-registered on the Open Science Framework: DOI 10.17605/OSF.IO/NFM27.We made several modifications to the protocol, with each change made prospectively in light of piloting. The changes to the protocol are described in the Supplementary Methods.

## Results

### Checklist

The items in the final checklist are presented in **Table 1**. The checklist contains 42 items across 8 domains. A glossary of MR technical terms in the checklist, adapted from the MR Dictionary [22], is presented in **Supplementary Table S1**.

The “Clear articulation of research question” domain has questions assessing whether authors clearly define the exposure(s), outcome(s), and number of hypothesises tested. “Data sources” examines reporting quality of the underlying GWASs used by each study including items relating to GWAS methods, such as quality control (QC), covariates included in models, measurement of traits, and recruitment of participants into studies. “2SMR specific assumptions” asks about reporting if the GWASs are representative of the same underlying population, and any quantification of the amount of sample overlap between GWASs.

“Data harmonisation” consists of two questions pertaining to data harmonisation, including how alleles were harmonised, and how palindromic SNPs were addressed. “Instrument construction” has various items related to how the instrument was constructed which includes questions on whether there was a clear description of how genetic variants were chosen, if the genetic variants were independent, how weights were estimated, the number of variants included in instruments, and the number of instruments used for each exposure. “Instrumental Variable (IV) assumptions and considerations” has questions about the instrumental variables assumptions, and asks how the three core assumptions were assessed, if additional sensitivity analyses were used, and whether any additional assumptions had been acknowledged.

The “Analytic methods” domain has miscellaneous questions about the reporting of other analytic methods, such as a definition of the primary model, power calculations, and the use of plots to visualise findings. “Reproducibility and open science” has two questions about “open science”: whether the data analysed has been provided or an explanation of where it can be accessed, and whether the R code used in the analysis has been presented.

### Included studies

The search resulted in 876 citations, including 505 unique studies. After full text screening, 87 studies were identified for inclusion in the review. Study selection is illustrated in **Figure 1**. Studies excluded in the full text screen and their reason for exclusion are presented in **Supplementary Table S3**. A list of studies included and excluded can be found **Supplementary Table S4** and **Supplementary Table S5**, respectively.

**Figure 1:**
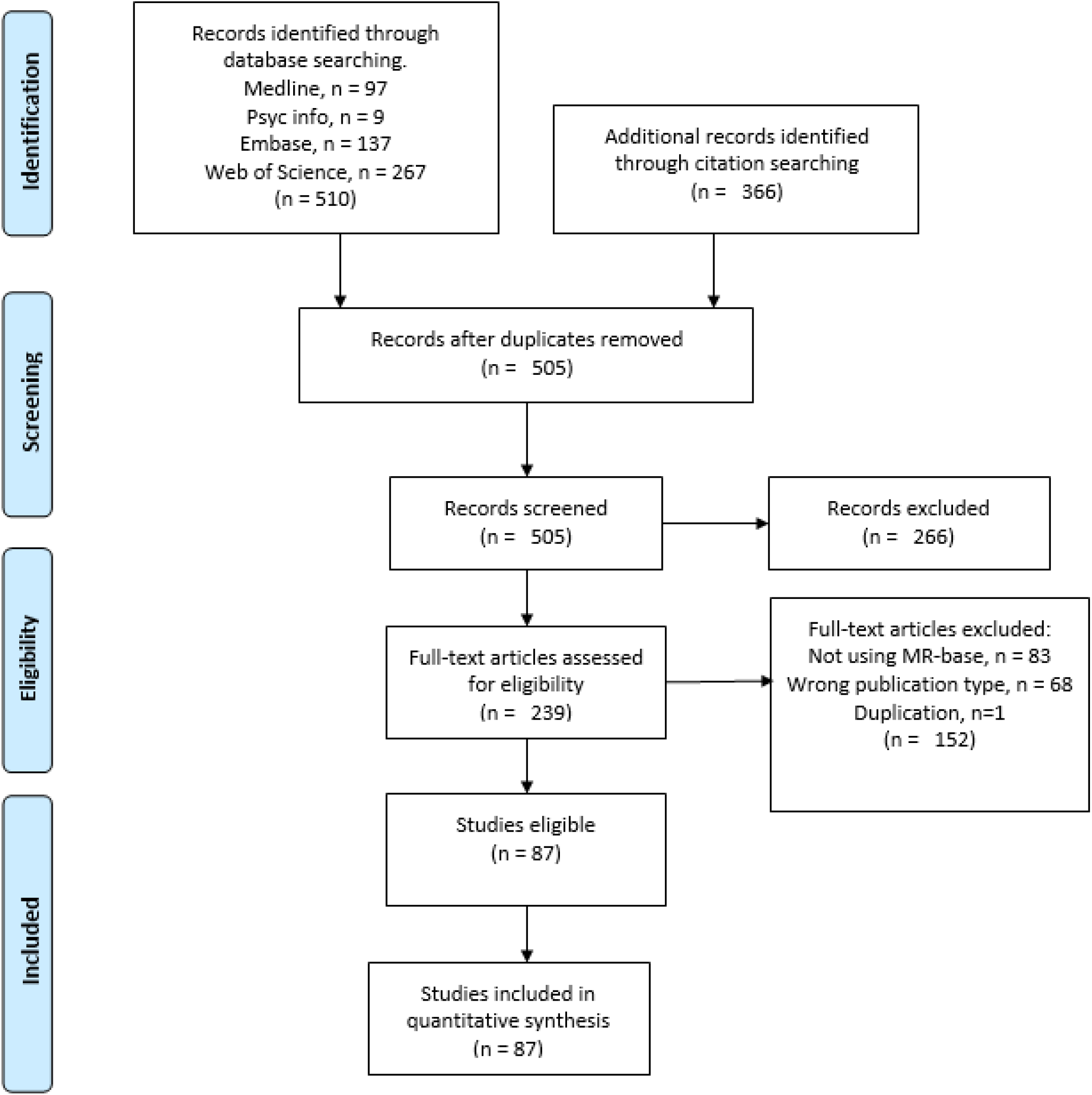
PRISMA Flow chart for study inclusion

Details of the data extracted and reviewers’ judgements on reporting quality are provided in **Supplementary Table S6**. 14 (16%) of the 87 studies, listed in **Supplementary Table S7**, were classified as ≥10 phenotype studies.

### Quality of reporting

**Table 2** presents the percentage of items reported per question, which ranged from 1% (item 28 -homogeneity/monotonicity/constant effect assumption) to 99% (item 31 – list primary model[s]). **Supplementary Figure S1** shows the number of studies that reported certain minimum thresholds for the percentage of items reported; 53 studies (61%) reported at least 50% of items, 5 (6%) reported at least 75% of items, and none reported 100% of items. The overall mean percentage of items reported in each study was 52% (SD = 14%).

**Table 2:** percentage of studies reporting each item, by item.

| Question | Total % items reported (n = 87*) | % in studies with ≥10 phenotypes (n = 14*) | % in studies with <10 phenotypes (n = 73*) |
| --- | --- | --- | --- |
| **Clear articulation of research question** |  |  |  |
| 1) Clearly define exposure(s) | 91.95 | 64.29 | 97.26 |
| 2) Clearly define outcome(s) | 90.80 | 71.43 | 94.52 |
| 3) State how many exposure/outcome relationships were tested in the main analysis | 91.95 | 85.71 | 93.15 |
| 4) Clearly state hypothesis under investigation | 89.66 | 78.57 | 91.78 |
| **Data sources** |  |  |  |
| 5) Provide an evidence trail to a description of the source GWAS** | 97.92 | 100.00 | 97.52 |
| 6) Describe the methods used to recruit participants into GWAS** | 16.15 | 13.88 | 16.56 |
| 7) State the number of participants included in GWAS** | 84.58 | 92.67 | 83.02 |
| 8) Describe how exposure and outcome were measured** | 37.62 | 40.47 | 37.07 |
| 9) State the units of measurement .** | 70.53 | 63.68 | 71.85 |
| 10) List covariates included in GWAS** | 27.55 | 26.38 | 27.77 |
| 11) Describe quality control procedures adopted in GWAS or explicitly provide a reference to it** | 24.67 | 29.42 | 23.76 |
| 12) If applicable, comment on any attempts made to address inadequate quality control or GWAS design in MR analysis** | 81.25 (of GWASs in 4 studies**) | 62.50 (of GWASs in 2 studies**) | 100.00 (of GWASs in 2 studies**) |
| 13) For MR studies examining where exposure and/or outcome is a disease/binary trait, describe if cases were incident or prevalent or both | 15.38 (10/65) | 9.09 (1/11) | 16.67 (9/54) |
| **2SMR specific assumptions** |  |  |  |
| 14) Provide adequate descriptive information on GWAS samples to assess whether they represent the same underlying population | 37.93 | 35.71 | 38.36 |
| 15) Provide information on sample overlap. | 33.33 | 35.71 | 32.88 |
| **Data harmonisation** |  |  |  |
| 16) Explain how reference alleles were harmonised across exposure and outcome datasets | 27.59 | 28.57 | 27.40 |
| 17) Describe how palindromic SNPs were addressed | 14.94 | 21.43 | 13.70 |
| **Instrument construction** |  |  |  |
| 18) Provide clear criteria or a clear description for the inclusion of genetic variants | 72.41 | 85.71 | 69.86 |
| 19) Describe whether genetic variant-exposure and genetic variant-outcome association estimates were obtained from a discovery | 66.67 | 57.14 | 68.49 |
| GWAS sample, replication GWAS sample, pooled discovery-replication GWAS samples, or a meta-analysis of two or more samples, or another source |  |  |  |
| 20) Describe whether the instrument was restricted to independent variants or whether the instrument consisted of correlated variants | 79.31 | 92.86 | 76.71 |
| 21) If the instrument consisted of independent variants, how was independence defined/ what were the clumping parameters used | 67.65 (46/68) | 78.57 (11/14) | 64.81 (35/54) |
| 22) If the instrument consisted of correlated variants, explain how this was accounted for in the model employed | 66.67 (4/6) | NA (n = 0) | 66.67 (4/6) |
| 23) State the number of primary instruments constructed | 97.70 | 92.86 | 98.63 |
| 24) State the number of included genetic variants. | 93.10 | 78.57 | 95.89 |
| **Instrumental Variable (IV) assumptions and considerations** |  |  |  |
| 25) Describe how IV assumption 1 (relevance) was assessed | 43.68 | 57.14 | 41.10 |
| 26) Describe how IV assumption 2 (independence) was assessed | 31.03 | 14.29 | 34.25 |
| 27) Describe how IV assumption 3 (exclusion restriction) was assessed | 88.51 | 78.57 | 90.41 |
| 28) Homogeneity/monotonicity/constant effect (IV assumption 4) was assessed | 1.15 | 0 | 1.37 |
| 29) If applicable, acknowledge all major assumptions introduced by falsification tests | 31.51 (23/73) | 16.67 (2/12) | 34.43 (21/61) |
| 30) Do they explain why the used sensitivity analyses were used | 90.90 (70/77) | 90.90 (10/11) | 90.91 (60/ 66) |
| **Analytic methods** |  |  |  |
| 31) List the primary model(s) employed to examine the exposure-outcome association | 98.85 | 92.86 | 100.00 |
| 32) Describe whether any proxy genetic variants were utilised in the analysis. | 24.14 | 21.43 | 24.66 |
| 33) Describe whether a power calculation has been performed. | 34.48 | 35.71 | 34.25 |
| 34) If there was >1 exposure/outcome relationships, either state whether this was corrected for multiple testing or provide justification for/against correcting for multiple testing (or discuss in discussion) | 50.94 (27/53) | 57.14 (8/14) | 48.72 (19/39) |
| 35) Describe any attempts to examine directionality of genetic variant-exposure and genetic variant-outcome associations | 39.08 | 42.86 | 38.36 |
| 36) If analyses were performed using a single genetic variant as an instrumental variable, was colocalisation performed? | 16.67 (1/6) | 50.00 (1/2) | 0 (0/4) |
| 37) If the exposure GWAS used a binary variable, is the causal effect described in terms | 29.89 | 21.43 | 31.51 |
| of liability (or susceptibility) to the exposure in the discussion. |  |  |  |
| 38) Do they describe the causal effect in terms of the units of measurement, or relate it to a clinically understandable scale of the exposure. | 42.31 (11/26) | 0 (0/5) | 52.38 (11/21) |
| 39) Describe whether any plots are presented to visualise results | 72.41 | 71.43 | 72.60 |
| 40) Describe whether other forms of MR specific bias could be present in analyses | 56.32 | 50.00 | 57.53 |
| **Reproducibility and open science** |  |  |  |
| 41) Present all data used to perform all analyses or describe where data can be accessed | 66.67 | 50.00 | 69.86 |
| 42) Provide R code for performing all analyses | 9.20 | 14.29 | 8.19 |

\* For items which were conditional, the percentages were calculated with respect to the number of eligible questions. The numbers in the brackets represent the numerator and denominator of the percentage.
\*\*Give information separately for exposure and outcome GWAS. Percentages represent the mean percentage of GWASs that reported the item in each study.

All items in the ‘Clear articulation of research question’ domain were well reported, with each item being reported in at least 90% of studies. In the ‘Data sources’ domain, the items requiring an evidence trail, sample size, and units were well reported (98%, 85%, and 71% respectively). In addition, around 81% of studies which stated a potential issue with the GWAS QC described a correction to address this issue. However, the other questions on study design details were less well reported, with 15-38% of studies reporting these items.

Neither question in the ‘2SMR specific assumption’ domain (on sample overlap and consistency of populations) was answered adequately, both being reported in fewer than 40% of studies. Likewise, the two questions in the ‘Data harmonisation’ domain were reported poorly, with fewer than 30% of studies providing information. All items in the ‘Instrument Construction’ domain were reported in most studies, with these items being reported at least 67% of the time. The worst reported items in this domain were the two conditional questions on details about the independence or non-independence of SNPs (both ∼67%), while the best reported items were the two on the number of instruments and genetic variants, both >93%. Items in the ‘Instrumental Variable (IV) assumptions and considerations’ domain were generally not well reported. 89% of studies stated the exclusion restriction assumption and why sensitivity analyses were performed. However, most studies did not describe the other assumptions (44% for relevance, and 31% for independence), or any assumptions of the falsification tests (32%).

Most studies described the primary model (99%), presented plots (72%), and described at least one MR specific limitation (56%). However, the other items in the ‘analytic methods’ domain were not reported by most studies. For example, just under 50% of studies which tested more than one hypothesis described the use of a multiple testing correction, and fewer than 25% described the conditions for using proxy variants. In the ‘Reproducibility and Open Science’ domain, 67% of studies presented all the data used, or information on how to access it, but fewer than 10% provided the analysis code.

### Multi-phenotype studies

Broadly, having < 10 or ≥ 10 phenotypes did not seem to impact reporting (overall mean percentage of items reported was 53% and 51%, respectively). However, studies with ≥ 10 phenotypes were less likely to clearly articulate the research question compared to studies with < 10 phenotypes. For example, 97% of studies with < 10 phenotypes clearly defined the exposure of interest as compared to only 64% of studies with ≥ 10 phenotypes. Likewise, 95% and 71% of these studies clearly defined the outcome(s) of interest, respectively.. On the other hand, studies with ≥ 10 phenotypes were more likely to provide sufficient information to assess the relevance assumption (57% vs 41%) and to describe a multiple testing correction when testing multiple hypotheses (57% vs 49%) (**Table 2**).

### Additional analyses

The mean reporting of checklist items did not differ markedly across the three years for which there were published studies, with a mean of 47% of items reported in 2017, 55% in 2018, and 51% in 2019 (**Supplementary Figure S2**). No studies were included from 2016. Additionally, items for which information is given to the user by MR-Base did appear to increase the probability that they were reported, with a mean reporting of 66% for items covered by both the R-package and web platform, 47% for items with partial coverage, and 36% for items not covered by either the R-package or web platform (**Supplementary Figure S3**). Studies that used the R Package reported more items (56%) than studies that did not (46%) **Supplementary Figure S4**).

The results of the additional review we conducted in a random sample of non-MR-Base studies can be found in the **Supplementary Results and Supplementary Table S8, S9, and S10**. Broadly, studies using MR-Base had a similar level of mean reporting (52%) to those that did not (47%, 95%CI: 34-60, **Figure 2**). Of the 11 items examined, only one, testing for directionality, implied a discrepancy between the MR-Base and non-MR-Base studies; with 39% of MR-Base studies exploring directionality, but only 4% (95%CI: 0 – 12) of non-MR-Base studies, and 6.7% (95%CI: 0 - 19) of non-MR-Base 2SMR studies.

**Figure 2:**
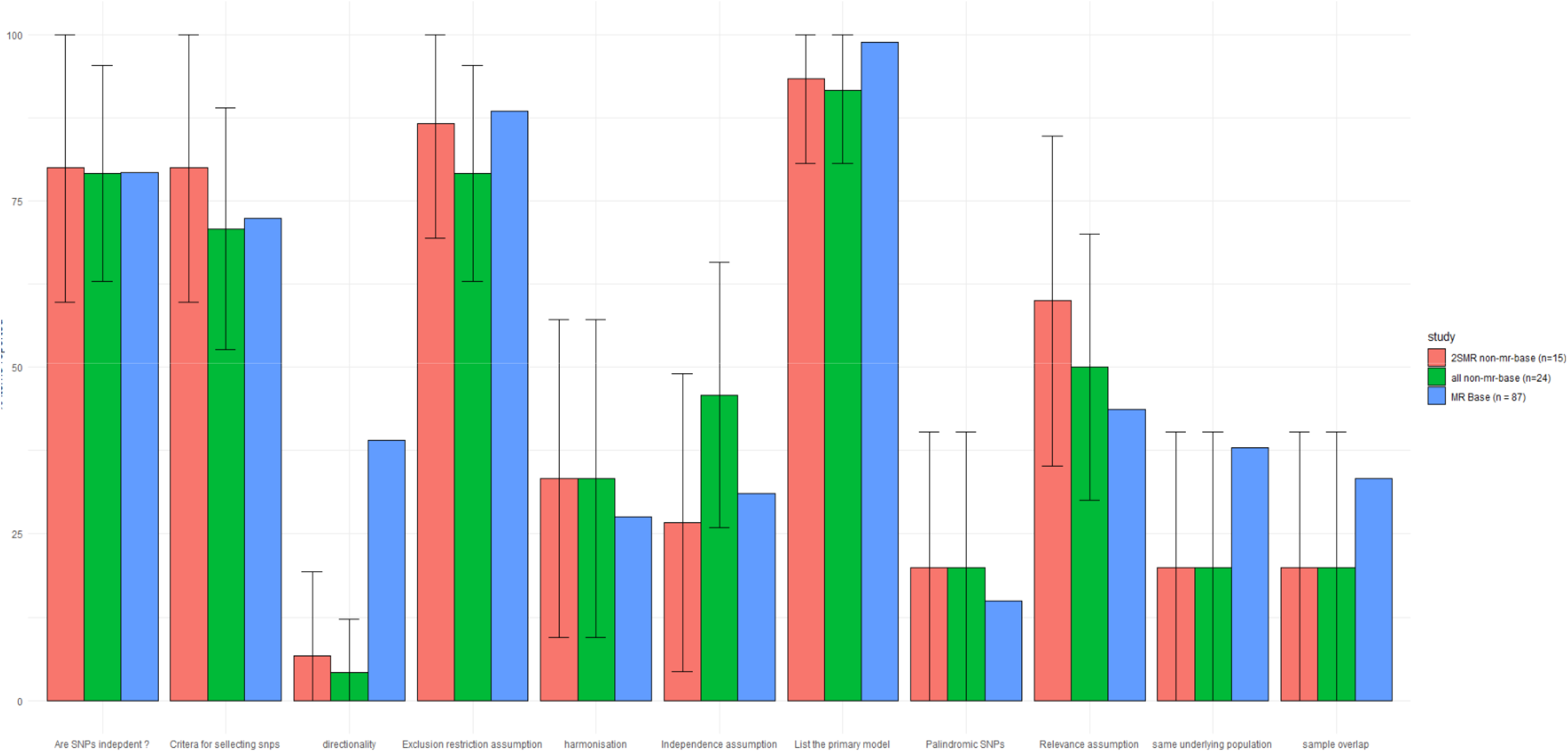
Comparison of MR-Base with non-MR-Base studies

## Discussion

### Summary of evidence

We sought to collate published studies that used the MR-Base platform to perform two sample Mendelian randomization (2SMR) analysis and to assess the quality of reporting of these studies. Our results reveal that reporting quality is generally poor, with 48% of items included in our reporting checklist not reported in an average study.

Many studies omitted information that is important for evaluating the methodological quality of a 2SMR study. This included, for example, information on the core instrumental variables assumptions (around 40% of studies had adequate reporting for the relevance assumption and around 30% had adequate reporting for the independence assumption), the assumptions of sensitivity analyses for the exclusion restriction assumption (around 30% of studies), and the 2SMR specific assumptions (< 40% of studies). Like any epidemiological design, MR can be affected by biases due to procedures employed during data collection[23]. However, papers included in this review did not tend to describe core aspects of the design and methods employed in the underlying GWAS used in MR analyses, including methods used to recruit participants into the study (16%), phenotypic measurements (38%), and quality control measures (25%). Importantly, these issues in reporting do not seem to be specific to MR-Base, with similar reporting quality being found in a random sample of MR papers which did not report using MR-Base (**Figure 2**).

Previous reviews of MR studies have found similarly poor levels of reporting. For example, Boef and colleagues found that only 33% of studies verified the F-statistic of the instruments, 70% provided any discussion of instrument-confounder associations, and 33% discussed the exclusion restriction assumption [24]. The increase in the number of studies in this review exploring the exclusion restriction assumption (88.5%) may be due to the development of several commonly used sensitivity analyses for appraising this assumption since the study by Boef and colleagues was published. MR-Base was developed around the time that a number of methods were starting to be developed to examine potential violations of the exclusion restriction assumption and part of the motivation for developing this platform was to improve the quality of causal inference by automating the application of exclusion restriction sensitivity analyses[16]. By contrast, the higher rate of studies assessing the independence assumption in Boef *et al* than this review (70% vs 31%) may be due to the higher prevalence of one-sample MR studies in Boef *at al*, since it is more straightforward to assess the confounding structure from individual-level data and because assessing this assumption did not feature in the reporting design of MR-Base. Similar to the poor levels of reporting of MR studies identified in Boef *et al*, in their review of 77 oncology MR studies, Lor *et al* found that only 48% stated the core MR assumptions, 31% estimated statistical power for analyses, 49% described the sample characteristics, and 62% assessed the relevance assumption (i.e. instrument-exposure correlation) [25].

Where MR-Base provides information pertaining to the relevant items of the checklist, this was linked to improvements in quality of reporting. However, our results do not necessarily imply that provision of information for item reporting by MR-Base improves reporting. The similarity of reporting with non-MR-Base studies implies that the apparent trend may be due to selection effects, with MR-Base providing information on items that are easier to report. However, this does not discount the possibility that integrating reporting nudges into the MR-Base platform, or other modifications, may improve reporting quality. For example, directionality testing is relatively unique to MR-Base compared to other MR software, and there was substantially higher reporting of this in MR-Base studies compared to non-MR-Base studies.

### Strengths and Limitations

There are several limitations to the current review. The included studies may not be a complete evaluation of all 2SMR papers that used MR-Base since our inclusion criteria required that authors explicitly mention MR-Base, the MR-Base R package, or cite one of the platform’s methods/description papers, or their iterations. It is likely that some studies will have been conducted using the platform or R package without providing any citation. If failure to report the statistical software used is indicative of poor reporting in general, then our study will overestimate the quality of reporting in studies using MR-Base. However, such studies would have been eligible for the non-MR-Base sample, and the consistency between these results implies that the difference in reporting may not be large.

Studies included in the review were published between 2016 and 2019. Based on the proportion of studies included, we estimate that approximately 155 relevant studies will have been published subsequently (**Supplementary Table S11**). However, no time trend in reporting quality was apparent in our data, so we consider it unlikely that more recent studies would differ markedly in their reporting quality as compared to those studies included in this review. However, if there were changes in reporting quality of studies between 2019 and 2021, this change could partially reflect an effect of the posting of the pre-print for the STROBE-MR checklist [21], made available in July 2019. Our final search date of April 2019 ensures that the quality of reporting presented is uncontaminated by the availability of the STROBE-MR guidelines. However, it will be important to evaluate the transparency of reporting in 2SMR before and after the publication of the STROBE-MR studies in a future study.

We excluded non-published pre-prints and other papers not published in peer-reviewed journals from this review to prevent bias that could arise from differences in methodological reporting across studies that underwent peer review (which could improve reporting) as compared to studies that did not. However, this also means the results of the study may not reflect the quality of reporting in preprints that used MR-Base. Additionally, this means that we were not able to evaluate whether peer review improves reporting quality of 2SMR studies that used MR-Base.

### Implications

Our review of the reporting quality of 87 two-sample Mendelian randomization studies conducted using MR-Base found that most studies were poorly well-reported. We make two sets of suggestions in light of this. First, MR-Base itself could be adapted to improve study reporting, for example by testing if the introduction of ‘nudges’ encouraging explicit thought of the analytic decisions being made enhances reporting quality. Future development of user interfaces according to agreed specifications of appropriate reporting may be effective in improving the quality of published papers. Second, we suggest that authors of all Mendelian randomization studies use, and that journals endorse, guidelines for reporting MR studies. Indeed, the development of this study’s checklist was used to inform the STROBE-MR reporting guidelines[21]. We hope that these guidelines will help to improve the quality of reporting of Mendelian randomization studies more generally, and would encourage its use by journals and researchers alike.

## Supporting information

Supplementary Results

## Data Availability

All data produced in the present work are contained in the manuscript

## Availability of data, code, and other materials

All materials used in this study are available in the supplementary materials or main text.

## Contributions

BW is the guarantor of the review. BW drafted the protocol and manuscript, all other authors commented on and edited these. RR, JY, KD contributed to the development of the selection criteria, the data extraction criteria and the search strategy. The search was implemented by BW and NC. BW, ND, RR, JY, and CMS conducted the data extraction. BW conducted the analysis, and BW, RR, and JY wrote the report. VS, RR, JY and BW implemented the search of non-MR-Base papers. All authors provided feedback on the final draft.

