## Supplementary Results for "Investigating the transparency of reporting in two-sample summary data Mendelian randomization studies"

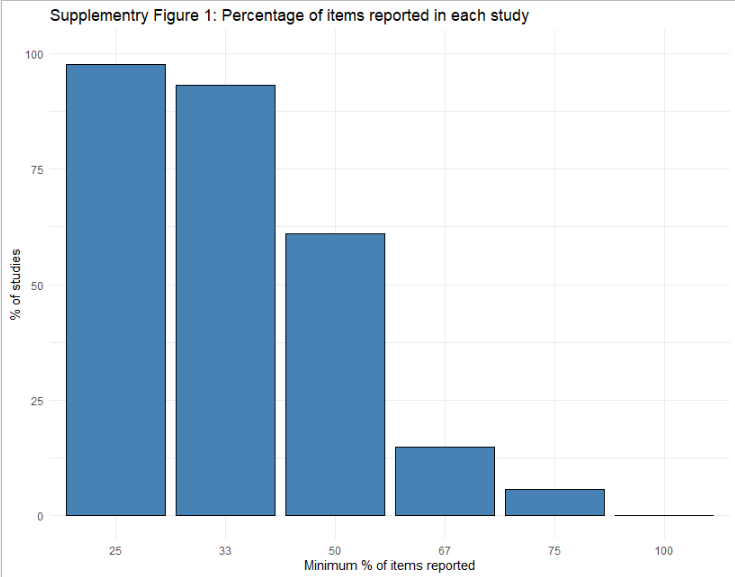

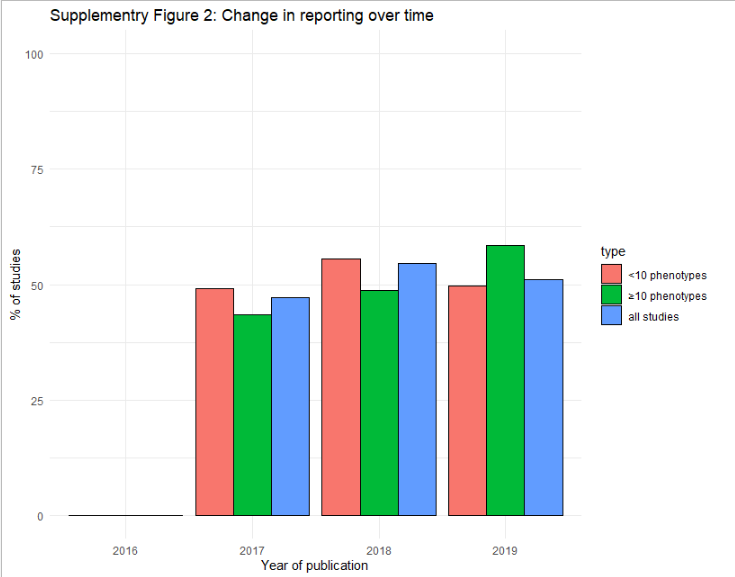

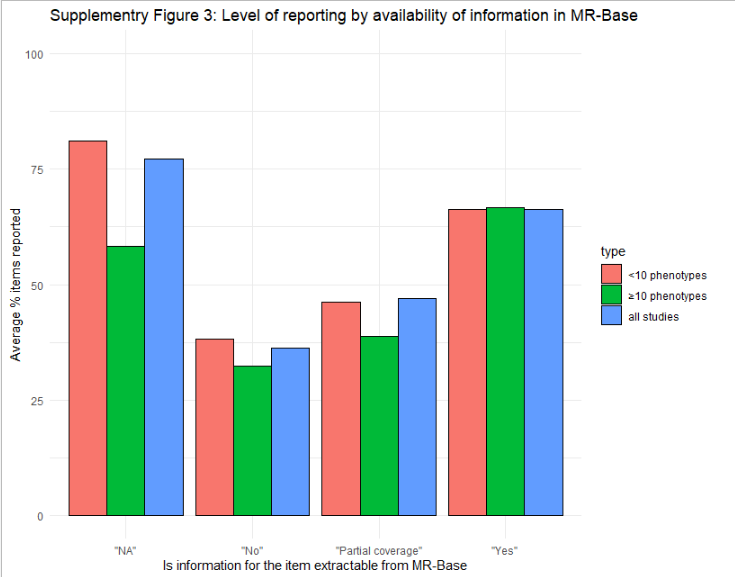

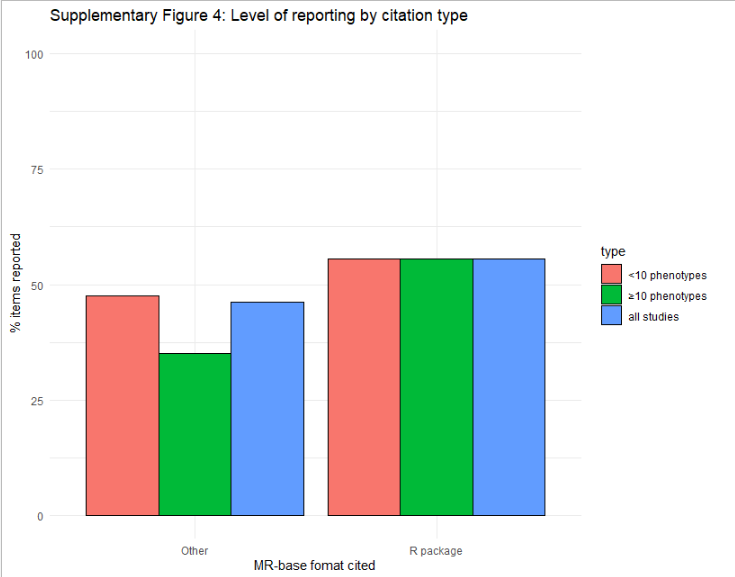

**Supplementary Table 3:** Studies excluded and reason for exclusion.

| Study | Reason for exclusion |
| --- | --- |
| Al-Dabhani 2018 | Wrong publication type |
| Anderson 2017 | Wrong publication type |
| Bartels 2018 | Wrong publication type |
| Bowden 2018 | Wrong study design |
| Brumpton 2019 | Wrong publication type |
| Budu-Aggrey 2018 | Wrong publication type |
| Budu-Aggrey 2019 | Wrong study design |
| Burgess 2017 | Wrong study design |
| Byrne 2017 | Wrong study design |
| Cao 2019 | Wrong study design |
| Caramaschi 2018 | Wrong study design |
| Carreras-Torres 2016 | Wrong study design |
| Carreras-Torres 2018 | Wrong study design |
| Carter 2018 | Wrong publication type |
| Carvalho 2019 | Wrong publication type |
| Chen 2018 | Wrong publication type |
| Cheung 2019 | Wrong study design |
| Cho 2018 | Wrong publication type |
| Corbin 2016 | Wrong study design |
| Crawford 2017 | Wrong publication type |
| Dardani 2018 | Wrong publication type |
| Disney-Hogg 2018 | Wrong study design |
| Ding 2018 | Wrong study design |
| Dixon 2019 | Wrong publication type |
| Dudding 2018 | Wrong study design |
| Ellervik 2019 | Wrong study design |
| Emanuelsson 2018 | Wrong study design |
| Evans 2019 | Wrong study design |
| Fang 2018 | Wrong study design |
| Fatima 2017 | Wrong publication type |
| Fluharty 2018 | Wrong publication type |
| Gage 2017 | Wrong study design |
| Gage 2016 | Wrong study design |
| Gauge 2018 | Wrong study design |
| Georgakis 2019 | Wrong study design |
| Gill 2018 | Wrong publication type |
| Gil 2018b | Wrong study design |
| Gill 2017 | Wrong study design |
| Gill 2018c | Wrong study design |
| Goto 2019 | Wrong study design |
| Grover 2018 | Wrong publication type |
| Gupta 2019 | Wrong study design |
| Hagenaars 2017 | Wrong publication type |
| Harshfield 2016 | Wrong publication type |
| Harati 2019 | Wrong publication type |
| Harati 2018 | Wrong publication type |
| Hartwig 2017 | Wrong study design |
| Hartiwg 2016 | Wrong study design |
| Hartwig 2016b | Wrong study design |
| Hartwig 2018 | Wrong publication type |
| Haworth 2017 | Wrong study design |
| Haycock 2017 | Wrong study design |
| Hemani 2017 | Wrong study design |
| Hillary 2019 | Wrong publication type |
| Holmes 2018 | Wrong study design |
| Howe 2019 | Wrong publication type |
| Howell 2018 | wrong study design |
| Huang 2017 | Wrong study design |
| Huang 2018 | Wrong study design |
| Hu 2019 | Wrong study design |
| Hypponen 2019 | Wrong publication type |
| Jones 2018 | Wrong publication type |
| Kar 2018 | Wrong publication type |
| Kia 2018 | Wrong study design |
| Kwok 2016 | Wrong study design |
| Lai 2018 | Wrong study deign |
| Lanktree 2018 | Wrong study design |
| Larsson 2017 | Wrong study design |
| Lee 2016 | Wrong study design |
| Li 2017 | Wrong publication type |
| Li 2018 | Wrong study design |
| Li 2018 | Wrong study design |
| Llano 2018 | Wrong publication type |
| Lloyd 2018 | Wrong publication type |
| Mack 2017 | Wrong study design |
| Magnus 2018 | Wrong publication type |
| McGowan 2018 | Wrong publication type |
| Meyer 2019 | Wrong publication type |
| Milaneschi 2019 | Wrong publication type |
| Mohammadi-Shemirani 2019 | Wrong study design |
| Mokry 2016 | Wrong study design |
| Morales 2016 | Wrong study design |
| Morris 2018 | Wrong publication type |
| Noordam 2017 | Wrong study design |
| Noordam 2018 | Wrong study design |
| Noyce 2017 | Wrong study design |
| Noyce 2018 | Wrong publication type |
| Noyce 2017b | Wrong publication type |
| Noyce 2019 | Wrong publication type |
| O’Connor 2017 | Wrong study design |
| Ogawa 2018 | Wrong publication type |
| Ong 2018 | Wrong study design |
| Pasman 2018 | Wrong publication type |
| Pasman 2018b | Wrong study design |
| Parisinos 2018 | Wrong study design |
| Parisinos 2018b | Wrong study design |
| Polimanti 2018 | Wrong publication type |
| Porcu 2019 | Wrong publication type |
| PRACTICAL consortium 2018 | Wrong study design |
| Prats-Uribe 2018 | Wrong publication type |
| Ravera 2018 | Wrong publication type |
| Richardson | Wrong study design |
| Richardson 2017 | Wrong study design |
| Richmond 2018 | Wrong publication type |
| Richmond 2017 | Wrong publication type |
| Rosoff 2019 | Wrong publication type |
| Sabater-Lleal 2019 | Wrong study design |
| Sallis 2019 | Wrong study design |
| Sandu 2018 | Wrong publication type |
| Schooling 2018 | Wrong publication type |
| So 2018 | Wrong publication type |
| Speed 2018 | Wrong publication type |
| Sun 2018 | Wrong study design |
| Sun 2018b | Wrong study design |
| Tokahashi 2019 | Wrong study design |
| Takahashi 2018 | Wrong study design |
| Than 2918 | Wrong publication type |
| Tan 2018b | Wrong publication type |
| Teurmer 2018 | Wrong study design |
| Timms 2018 | Wrong publication type |
| Trajanoska 2018 | Wrong study design |
| Valdes-Marquez 2019 | Wrong study deigns |
| Van Vliet 2018 | Wrong study design |
| Verweij 2018 | Wrong study design |
| Walker 2018 | Wrong publication type |
| Walker 2018b | Wrong study design |
| Wang 2018 | Wrong publication type |
| Wang 2019 | Wrong study design |
| Ward-Caviness 2018 | Wrong publication type |
| Ware 2017 | Wrong study design |
| Wendt 2018 | Wrong publication type |
| Weng 2018 | Wrong study design |
| Wiklund 2018 | Wrong publication type |
| Wood 2019 | Wrong study design |
| Wu 2016 | Wrong study design |
| Xu 2017 | Wrong study design |
| Xu 2017b | Wrong study design |
| Yarmolinksy 2018 | Wrong publication type |
| Yin 2017 | Wrong study design |
| Zen 2018 | Wrong publication type |
| Zhan 2017 | Wrong study design |
| Zhao 2018 | Wrong study design |
| Zheng 2018 | Wrong publication type |
| Zheng 2016 | Wrong publication type |
| Zhuang 2019 | Wrong study design |
| Richardson 2017 | Wrong study design |

**Supplementary Table 4:** Citations of studies included.

Adams CD, Richmond R, Ferreira DLS, Spiller W, Tan V, Zheng J, et al. Circulating Metabolic Biomarkers of Screen-Detected Prostate Cancer in the ProtecT Study. Cancer Epidemiol Biomarkers Prev. 2019;28(1):208 doi: 10.1158/1055-9965.EPI-18-0079.

Alghamdi J., Matou-Nasri S., Alghamdi F., Alghamdi S., Alfadhel M., Padmanabhan S. Risk of neuropsychiatric adverse effects of lipid-lowering drugs: A mendelian randomization study. Int J Neuropsychopharmacol. 2018;21(12):1067–75.  doi: 10.1093/ijnp/pyy060.

Arathimos R., Granell R., Haycock P., Richmond R.C., Yarmolinsky J., Relton C.L., et al. Genetic and observational evidence supports a causal role of sex hormones on the development of asthma. Thorax [Internet]. 2019; Available from: <http://thorax.bmj.com/>

Arathimos R, Suderman M, Sharp GC, Burrows K, Granell R, Tilling K, et al. Epigenome-wide association study of asthma and wheeze in childhood and adolescence. Clinical Epigenetics. 2017;9(1):112. doi: 10.1186/s13148-017-0414-7

Au Yeung SL, Borges M-C, Lawlor DA. Association of Genetic Instrumental Variables for Lung Function on Coronary Artery Disease Risk: A 2-Sample Mendelian Randomization Study. Circ Genom Precis Med. 2018;11(4):e001952. <https://doi.org/10.1161/CIRCGEN.117.001952>

Au Yeung SL, Lam HSHS, Schooling CM. Vascular Endothelial Growth Factor and Ischemic Heart Disease Risk: A Mendelian Randomization Study. Journal of the American Heart Association [Internet]. 2017;6(8). Available from: <https://www.ahajournals.org/doi/10.1161/JAHA.117.005619>

Aziz N, Weydt P, Aziz NA, Weydt P. Telomere length as a modifier of age-at-onset in Huntington disease: a        two-sample Mendelian randomization study. JOURNAL OF NEUROLOGY. 2018;265(9):2149–51. doi: 10.1007/s00415-018-8972-y.

Bae S.-.C., Lee Y.H. Alcohol intake and risk of rheumatoid arthritis: a Mendelian randomization study. Z Rheumatol [Internet]. 2018; Available from: <http://www.steinkopff.springer.de/393/home.htm>

Bae S.-C., Lee Y.H. Causal association between rheumatoid arthritis and a decreased risk of Alzheimer’s disease: A Mendelian randomization study. Z Rheumatol. 2018;1–6. doi: 10.1007/s00393-018-0504-8.

Bae S, Lee Y, Bae SC, Lee YH. Alcohol intake and risk of systemic lupus erythematosus: a Mendelian        randomization study. LUPUS. 2019;28(2):174–80. [https://doi.org/10.1177/0961203318817832](https://doi.org/10.1177%2F0961203318817832)

Bae S, Lee Y, Bae S-C, Lee YH. Coffee consumption and the risk of rheumatoid arthritis and systemic        lupus erythematosus: a Mendelian randomization study. CLINICAL RHEUMATOLOGY. 2018;37(10):2875–9. DOI: [10.1007/s10067-018-4278-9](https://doi.org/10.1007/s10067-018-4278-9)

Bae S, Lee Y, Bae S-C, Lee YH. Vitamin D level and risk of systemic lupus erythematosus and rheumatoid        arthritis: a Mendelian randomization. CLINICAL RHEUMATOLOGY. 2018;37(9):2415–21.  DOI: [10.1007/s10067-018-4152-9](https://doi.org/10.1007/s10067-018-4152-9)

Bae S, Lee Y, Bae S-C, Lee YH. Causal association between body mass index and risk of rheumatoid        arthritis: A Mendelian randomization study. EUROPEAN JOURNAL OF CLINICAL INVESTIGATION. 2019;49(4).  DOI: [10.1111/eci.13076](https://doi.org/10.1111/eci.13076)

Bandres-Ciga S, Noyce AJ, Hemani G, Nicolas A, Calvo A, Mora G, et al. Shared polygenic risk and causal inferences in amyotrophic lateral sclerosis. Ann Neurol. 2019;85(4):470–81.  DOI: [10.1002/ana.25431](https://doi.org/10.1002/ana.25431)

Bell JA, Carslake D, Wade KH, Richmond RC, Langdon RJ, Vincent EE, et al. Influence of puberty timing on adiposity and cardiometabolic traits: A Mendelian randomisation study. PLOS Medicine. 2018;15(8):e1002641. <https://doi.org/10.1371/journal.pmed.1002641>

Bovijn J, Jackson L, Censin J, Chen C-Y, Laisk T, Laber S, et al. GWAS Identifies Risk Locus for Erectile Dysfunction and Implicates Hypothalamic Neurobiology and Diabetes in Etiology. The American Journal of Human Genetics. 2019;104(1):157–63.  DOI: [10.1016/j.ajhg.2018.11.004](https://doi.org/10.1016/j.ajhg.2018.11.004)

Brower M.A., Hai Y., Jones M.R., Guo X., Chen Y.-D.I., Rotter J.I., et al. Bidirectional Mendelian randomization to explore the causal relationships between body mass index and polycystic ovary syndrome. Hum Reprod. 2019;34(1):127–36. DOI: [10.1093/humrep/dey343](https://doi.org/10.1093/humrep/dey343)

Brumpton BM, Fritsche LG, Zheng J, Nielsen JB, Mannila M, Surakka I, et al. Variation in Serum PCSK9 (Proprotein Convertase Subtilisin/Kexin Type 9), Cardiovascular Disease Risk, and an Investigation of Potential Unanticipated Effects of PCSK9 Inhibition: A Genome-Wide Association Study and Mendelian Randomization Study in the HUNT, Norway. Circulation: Genomic and Precision Medicine [Internet]. 2019;12(1). Available from: <https://www.ahajournals.org/doi/10.1161/CIRCGEN.118.002335>

Caramaschi D, Sharp GC, Nohr EA, Berryman K, Lewis SJ, Davey Smith G, et al. Exploring a causal role of DNA methylation in the relationship between maternal vitamin B12 during pregnancy and child’s IQ at age 8, cognitive performance and educational attainment: a two-step Mendelian randomization study. Human Molecular Genetics. 2017;26(15):3001–13. DOI: [10.1093/hmg/ddx164](https://doi.org/10.1093/hmg/ddx164)

Carreras-Torres R, Johansson M, Haycock PC, Wade KH, Relton CL, Martin RM, et al. Obesity, metabolic factors and risk of different histological types of lung cancer: A Mendelian randomization study. PLOS ONE. 2017;12(6):e0177875. <https://doi.org/10.1371/journal.pone.0177875>

Cecil CAM, Walton E, Pingault J-B, Provençal N, Pappa I, Vitaro F, et al. DRD4 methylation as a potential biomarker for physical aggression: An epigenome-wide, cross-tissue investigation. American Journal of Medical Genetics Part B: Neuropsychiatric Genetics. 2018;177(8):746–64. doi: 10.1002/ajmg.b.32689

Cheng W-W, Zhu Q, Zhang H-Y. Mineral Nutrition and the Risk of Chronic Diseases: A Mendelian Randomization Study. Nutrients. 2019;11(2):378. doi: 10.3390/nu11020378.

Cherny SS, Freidin MB, Williams FMK, Livshits G, Güerri-Fernández R. The analysis of causal relationships between blood lipid levels and BMD. PLOS ONE. 2019;14(2):e0212464. https://doi.org/ 10.1371/journal.pone.0212464

Choi K.W., Chen C.-Y., Stein M.B., Klimentidis Y.C., Wang M.-J., Koenen K.C., et al. Assessment of Bidirectional Relationships between Physical Activity and Depression among Adults: A 2-Sample Mendelian Randomization Study. JAMA Psychiatry [Internet]. 2019; Available from: <http://archpsyc.jamanetwork.com/issues.aspx>

Colodro-Conde L, Couvy-Duchesne B, Whitfield JB, Streit F, Gordon S, Kemper KE, et al. Association Between Population Density and Genetic Risk for Schizophrenia. JAMA Psychiatry. 2018;75(9):901–10. doi: 10.1001/jamapsychiatry.2018.1581.

Cousminer D, Mitchell J, Chesi A, Roy S, Kalkwarf H, Lappe J, et al. Genetically Determined Later Puberty Impacts Lowered Bone Mineral        Density in Childhood and Adulthood. JOURNAL OF BONE AND MINERAL RESEARCH. 2018;33(3):430–6.  doi: 10.1002/jbmr.3320.

Dashti H.S., Redline S., Saxena R. Polygenic risk score identifies associations between sleep duration and diseases determined from an electronic medical record biobank. Sleep. 2019;42(3):zsy247. doi: 10.1093/sleep/zsy247.

Dashti HS, Jones SE, Wood AR, Lane JM, van Hees VT, Wang H, et al. Genome-wide association study identifies genetic loci for self-reported habitual sleep duration supported by accelerometer-derived estimates. Nat Commun. 2019;10(1):1100. doi: 10.1038/s41467-019-08917-4.

Dimitrakopoulou V, Tsilidis K, Haycock P, Dimou N, Al-Dabhani K, Martin R, et al. Circulating vitamin D concentration and risk of seven cancers: Mendelian        randomisation study. BMJ-BRITISH MEDICAL JOURNAL. 2017;359. doi: 10.1136/bmj.j4761.

Doherty A, Smith-Byrne K, Ferreira T, Holmes M, Holmes C, Pulit S, et al. GWAS identifies 14 loci for device-measured physical activity and sleep        duration. NATURE COMMUNICATIONS. 2018;9. doi: 10.1038/s41467-018-07743-4.

Fatima T, McKinney C, Major T, Stamp L, Dalbeth N, Iverson C, et al. The relationship between ferritin and urate levels and risk of gout. ARTHRITIS RESEARCH & THERAPY. 2018;20.

Gibson M, Munafo MR, Taylor A, Treur JL. Evidence for genetic correlations and bidirectional, causal effects between smoking and sleep behaviours [Internet]. Genetics; 2018. Available from: <http://biorxiv.org/lookup/doi/10.1101/258384>

Grace C, Clarke R, Goel A, Farrall M, Watkins H, Hopewell J, et al. Lack of genetic support for shared aetiology of Coronary Artery Disease        and Late-onset Alzheimer’s disease. SCIENTIFIC REPORTS. 2018;8. https://doi.org/10.1038/s41598-018-25460-2

Haas ME, Aragam KG, Emdin CA, Bick AG, Hemani G, Smith GD, Kathiresan S, International Consortium for Blood Pressure. Genetic association of albuminuria with cardiometabolic disease and blood pressure. The American Journal of Human Genetics. 2018 Oct 4;103(4):461-73. doi: 10.1016/j.ajhg.2018.08.004.

Havdahl A, Mitchell R, Paternoster L, Smith G, Havdahl A, Mitchell R, et al. Investigating causality in the association between vitamin D status and        self-reported tiredness. SCIENTIFIC REPORTS. 2019;9.  https://doi.org/10.1038/s41598-019-39359-z

Hemani G, Tilling K, Davey Smith G. Orienting The Causal Relationship Between Imprecisely Measured Traits Using Genetic Instruments [Internet]. Systems Biology; 2017. Available from: <http://biorxiv.org/lookup/doi/10.1101/117101>

Howe LJ, Richardson TG, Arathimos R, Alvizi L, Passos-Bueno MR, Stanier P, et al. Evidence for DNA methylation mediating genetic liability to non-syndromic cleft lip/palate. Epigenomics. 2019;11(2):133–45. <https://doi.org/10.2217/epi-2018-0091>

Howe LJ, Lee MK, Sharp GC, Smith GD, Pourcain BS, Shaffer JR, et al. Investigating the shared genetics of non-syndromic cleft lip/palate and facial morphology. PLOS Genetics. 2018;14(8):e1007501. <https://doi.org/10.1371/journal.pgen.1007501>

Johansson M., Carreras-Torres R., Scelo G., Purdue M.P., Mariosa D., Muller D.C., et al. The influence of obesity-related factors in the etiology of renal cell carcinoma-A mendelian randomization study. PLoS Med. 2019;16(1):e1002724. doi: 10.1371/journal.pmed.1002724.

Jones SE, Lane JM, Wood AR, Hees VT van, Tyrrell J, Beaumont RN, et al. Genome-wide association analyses of chronotype in 697,828 individuals provides insights into circadian rhythms. Nature Communications. 2019;10(1):343. doi: 10.1038/s41467-018-08259-7.

Joshi P, Pirastu N, Kentistou K, Fischer K, Hofer E, Schraut K, et al. Genome-wide meta-analysis associates HLA-DQA1/DRB1 and LPA and lifestyle        factors with human longevity. NATURE COMMUNICATIONS. 2017;8.  doi: 10.1038/s41467-017-00934-5

Kar S.P., Andrulis I.L., Brenner H., Burgess S., Chang-Claude J., Considine D., et al. The association between weight at birth and breast cancer risk revisited using Mendelian randomisation. Eur J Epidemiol [Internet]. 2019; Available from: <http://www.wkap.nl/journalhome.htm/0393-2990>

Kar SP, Brenner H, Giles GG, Huo D, Milne RL, Rennert G, et al. Body mass index and the association between low-density lipoprotein cholesterol as predicted by HMGCR genetic variants and breast cancer risk. International Journal of Epidemiology [Internet]. 2019; Available from: <https://academic.oup.com/ije/advance-article/doi/10.1093/ije/dyz047/5423849>

Keefe JA, Hwang S-J, Huan T, Mendelson M, Yao C, Courchesne P, et al. Evidence for a Causal Role of the SH2B3 -β 2 M Axis in Blood Pressure Regulation: Framingham Heart Study. Hypertension. 2019;73(2):497–503. h[ttps://doi.org/10.1161/HYPERTENSIONAHA.118.12094](https://doi.org/10.1161/HYPERTENSIONAHA.118.12094)

Kho P, Glubb D, Thompson D, Spurdle A, O’Mara T, Kho PF, et al. Assessing the Role of Selenium in Endometrial Cancer Risk: A Mendelian        Randomization Study. FRONTIERS IN ONCOLOGY. 2019;9. doi: 10.3389/fonc.2019.00182.

Lane J.M., Jones S., Dashti H.S., Wood A., Van Hees V., Spiegelhalder K., et al. Biological and clinical insights from genetics of insomnia symptoms. Sleep. 2018;41:A7. doi: 10.1038/s41588-019-0361-7.

Lee Y.H. Causal association between smoking behavior and the decreased risk of osteoarthritis: a Mendelian randomization. Z Rheumatol. 2018;1–6. doi: 10.1007/s00393-018-0505-7.

Lee Y.H. Gout and the risk of Alzheimer’s disease: A Mendelian randomization study. Int J Rheum Dis [Internet]. 2019; Available from: <http://onlinelibrary.wiley.com/journal/10.1111/(ISSN)1756-185X>

Lee Y, Lee YH. Assessing the causal association between smoking behavior and risk of        gout using a Mendelian randomization study. CLINICAL RHEUMATOLOGY. 2018;37(11):3099–105. doi: 10.1007/s10067-018-4210-3.

Lee Y, Lee YH. Investigating the possible causal association of coffee consumption with        osteoarthritis risk using a Mendelian randomization analysis. CLINICAL RHEUMATOLOGY. 2018;37(11):3133–9. doi: 10.1007/s10067-018-4252-6.

Lee YH, Song GG. Causal Association between Rheumatoid Arthritis with the Increased Risk of Type 2 Diabetes: A Mendelian Randomization Analysis. Journal of Rheumatic Diseases. 2019;26(2):131. <http://orcid.org/0000-0003-4213-1909>

Li Z, Chen P, Chen J, Xu Y, Wang Q, Li X, et al. Glucose and Insulin-Related Traits, Type 2 Diabetes and Risk of Schizophrenia: A Mendelian Randomization Study. EBioMedicine. 2018;34:182–8. doi: 10.1016/j.ebiom.2018.07.037.

Ligthart S, Vaez A, Võsa U, Stathopoulou MG, de Vries PS, Prins BP, et al. Genome Analyses of >200,000 Individuals Identify 58 Loci for Chronic Inflammation and Highlight Pathways that Link Inflammation and Complex Disorders. The American Journal of Human Genetics. 2018;103(5):691–706. doi: 10.1016/j.ajhg.2018.09.009.

Liyanage UE, Law MH, Ong JS, Cust AE, Mann GJ, Ward SV, et al. Polyunsaturated fatty acids and risk of melanoma: A Mendelian randomisation analysis: PUFAs and risk of melanoma. International Journal of Cancer. 2018;143(3):508–14. doi: 10.1002/ijc.31334.

Liyanage UE, Ong J-S, An J, Gharahkhani P, Law MH, MacGregor S. Mendelian randomization study for genetically predicted polyunsaturated fatty acids levels on overall cancer risk and mortality. Cancer Epidemiology Biomarkers & Prevention. 2019;cebp.0940.2018. doi: 10.1158/1055-9965.EPI-18-0940.

Luo S., Au Yeung S.L., Zhao J.V., Burgess S., Schooling C.M. Association of genetically predicted testosterone with thromboembolism, heart failure, and myocardial infarction: Mendelian randomisation study in UK Biobank. BMJ (Online). 2019;364:l476. doi: <https://doi.org/10.1136/bmj.l476>

Moen G-H, Qvigstad E, Birkeland KI, Evans DM, Sommer C. Are serum concentrations of vitamin B-12 causally related to cardiometabolic risk factors and disease? A Mendelian randomization study. Am J Clin Nutr. 2018;108(2):398–404. doi: 10.1093/ajcn/nqy101.

Morgan RA, Beck KR, Nixon M, Homer NZM, Crawford AA, Melchers D, et al. Carbonyl reductase 1 catalyzes 20β-reduction of glucocorticoids, modulating receptor activation and metabolic complications of obesity. Scientific Reports. 2017;7(1):10633. doi: 10.1038/s41598-017-10410-1.

Morris AP, Le TH, Wu H, Akbarov A, van der Most PJ, Hemani G, et al. Trans-ethnic kidney function association study reveals putative causal genes and effects on kidney-specific disease aetiologies. Nature Communications. 2019;10(1):29. doi: 10.1038/s41467-018-07867-7.

Mountjoy E, Davies N, Plotnikov D, Smith G, Rodriguez S, Williams C, et al. Education and myopia: assessing the direction of causality by mendelian        randomisation. BMJ-BRITISH MEDICAL JOURNAL. 2018;361. doi: <https://doi.org/10.1136/bmj.k2022>

Nowake C, Arnlov J, Nowake C, Arnlov J. A Mendelian randomization study of the effects of blood lipids on breast        cancer risk. NATURE COMMUNICATIONS. 2018;9. doi: 10.1038/s41467-018-06467-9.

Ogawa K, Stuart PE, Tsoi LC, Suzuki K, Nair RP, Mochizuki H, et al. A Transethnic Mendelian Randomization Study Identifies Causality of Obesity on Risk of Psoriasis. Journal of Investigative Dermatology. 2018;S0022202X18329154. doi: 10.1016/j.jid.2018.11.023.

Ong J-S, An J, Law MH, Whiteman DC, Neale RE, Gharahkhani P, et al. Height and overall cancer risk and mortality: evidence from a Mendelian randomisation study on 310,000 UK Biobank participants. British Journal of Cancer. 2018;118(9):1262–7. doi: 10.1038/s41416-018-0063-4.

Pasman JA, Verweij KJH, Gerring Z, Stringer S, Sanchez-Roige S, Treur JL, et al. GWAS of lifetime cannabis use reveals new risk loci, genetic overlap with psychiatric traits, and a causal influence of schizophrenia. Nat Neurosci. 2018;21(9):1161–70. doi: 10.1038/s41593-018-0206-1

PGC-ED, ENIGMA Genetics Working Group, Walton E, Hibar D, Yilmaz Z, Jahanshad N, et al. Exploration of Shared Genetic Architecture Between Subcortical Brain Volumes and Anorexia Nervosa. Molecular Neurobiology [Internet]. 2018; Available from: <http://link.springer.com/10.1007/s12035-018-1439-4>

Ravera S., Carrasco N., Gelernter J., Polimanti R. Phenomic impact of genetically-determined euthyroid function and molecular differences between thyroid disorders. J Clin Med. 2018;7(10):296.  <https://doi.org/10.3390/jcm7100296>

Reed Z, Micali N, Bulik C, Smith G, Wade K,. Assessing the causal role of adiposity on disordered eating in        childhood, adolescence, and adulthood: a Mendelian randomization        analysis. AMERICAN JOURNAL OF CLINICAL NUTRITION. 2017;106(3):764–72.  <https://doi.org/10.3945/ajcn.117.154104>

Richardson T.G., Haycock P.C., Zheng J., Timpson N.J., Gaunt T.R., Smith G.D., et al. Systematic Mendelian randomization framework elucidates hundreds of CpG sites which may mediate the influence of genetic variants on disease. Hum Mol Genet. 2018;27(18):3293–304. doi: 10.1093/hmg/ddy210.

Shadrina AS, Sharapov SZ, Shashkova TI, Tsepilov YA. Varicose veins of lower extremities: insights from the first large-scale genetic study [Internet]. Genomics; 2018. Available from: <http://biorxiv.org/lookup/doi/10.1101/368365>

Shi J, Wu L, Zheng W, Wen W, Wang S, Shu X, et al. Genetic Evidence for the Association between Schizophrenia and Breast Cancer. J Psychiatr Brain Sci. 2018;3(4). doi: 10.20900/jpbs.20180007.

Silbernagel G, Scharnagl H, Kleber M, Delgado G, Stojakovic T, Laaksonen R, et al. LDL triglycerides, hepatic lipase activity, and coronary artery disease:        An epidemiologic and Mendelian randomization study. ATHEROSCLEROSIS. 2019;282:37–44. doi: 10.1016/j.atherosclerosis.2018.12.024.

So H-C, Chau K-L, Ao F-K, Mo C-H, Sham P-C. Exploring shared genetic bases and causal relationships of schizophrenia and bipolar disorder with 28 cardiovascular and metabolic traits. Psychological Medicine. 2018;1–13. doi: 10.1017/S0033291718001812.

Soler Artigas M., Sanchez-Mora C., Rovira P., Richarte V., Garcia-Martinez I., Pagerols M., et al. Attention-deficit/hyperactivity disorder and lifetime cannabis use: genetic overlap and causality. Mol Psychiatry [Internet]. 2019; Available from: <http://www.nature.com/mp/index.html>

Song GG, Lee YH. Causal Association between Bone Mineral Density and Osteoarthritis: A Mendelian Randomization Study. Journal of Rheumatic Diseases. 2019;26(2):104. http://www.jrd.or.kr/journal/view.html?doi=10.4078/jrd.2019.26.2.104

Speed D, Hemani G, Speed MS, Major Depressive Disorder Working Group of the Psychiatric Genomics Consortium, Børglum AD, Østergaard SD. Investigating the causal relationship between neuroticism and depression via Mendelian randomization. Acta Psychiatrica Scandinavica. 2019;139(4):395–7.  <https://doi.org/10.1111/acps.13009>

Tan VY, Biernacka KM, Dudding T, Bonilla C, Gilbert R, Kaplan RC, et al. Reassessing the Association between Circulating Vitamin D and IGFBP-3: Observational and Mendelian Randomization Estimates from Independent Sources. Cancer Epidemiology Biomarkers & Prevention. 2018;27(12):1462–71. doi: 10.1158/1055-9965.EPI-18-0113.

Taylor K, Davey Smith G, Relton CL, Gaunt TR, Richardson TG. Prioritizing putative influential genes in cardiovascular disease susceptibility by applying tissue-specific Mendelian randomization. Genome Med. 2019;11(1):6. doi: 10.1186/s13073-019-0613-2.

Tikkanen E, Gustafsson S, Amar D, Shcherbina A, Waggott D, Ashley EA, et al. Biological Insights Into Muscular Strength: Genetic Findings in the UK Biobank. Scientific Reports. 2018;8(1):6451. doi: 10.1038/s41598-018-24735-y.

Treur JL, Gibson M, Taylor AE, Rogers PJ, Munafò MR. Investigating genetic correlations and causal effects between caffeine consumption and sleep behaviours. Journal of Sleep Research. 2018;27(5):e12695. doi: 10.1111/jsr.12695.

Williams D, Hagg S, Pedersen N, Williams DM, Hagg S, Pedersen NL. Circulating antioxidants and Alzheimer disease prevention: a Mendelian        randomization study. AMERICAN JOURNAL OF CLINICAL NUTRITION. 2019;109(1):90–8. doi: 10.1093/ajcn/nqy225.

Williams DM, Karlsson IK, Pedersen NL, Hägg S. Circulating insulin-like growth factors and Alzheimer disease: A mendelian randomization study. Neurology. 2018;90(4):e291–7. doi: 10.1212/WNL.0000000000004854.

Wootton RE, Lawn RB, Millard LAC, Davies NM, Taylor AE, Munafò MR, et al. Evaluation of the causal effects between subjective wellbeing and cardiometabolic health: mendelian randomisation study. BMJ. 2018;362:k3788. doi: 10.1136/bmj.k3788.

Yao C, Chen G, Song C, Keefe J, Mendelson M, Huan T, et al. Genome‐wide mapping of plasma protein QTLs identifies putatively causal genes and pathways for cardiovascular disease. Naturez Communications. 2018;9(1):3268. doi: 10.1038/s41467-018-05512-x.

Yeung CHC, Au Yeung SL, Fong SSM, Schooling CM. Lean mass, grip strength and risk of type 2 diabetes: a bi-directional Mendelian randomisation study. Diabetologia. 2019;62(5):789–99. doi: 10.1007/s00125-019-4826-0.

Zanetti D, Tikkanen E, Gustafsson S, Priest JR, Burgess S, Ingelsson E. Birthweight, Type 2 Diabetes Mellitus, and Cardiovascular Disease: Addressing the Barker Hypothesis With Mendelian Randomization. Circulation: Genomic and Precision Medicine [Internet]. 2018;11(6). Available from: <https://www.ahajournals.org/doi/10.1161/CIRCGEN.117.002054>

Zhang Q, Greenbaum J, Zhang W-D, Sun C-Q, Deng H-W. Age at menarche and osteoporosis: A Mendelian randomization study. Bone. 2018;117:91–7. doi: 10.1016/j.bone.2018.09.015

Zhang X, Lv W, Qiu B, Zhang L, Qin J, Tang F, et al. Assessing causal estimates of the association of obesity-related traits        with coronary artery disease using a Mendelian randomization approach. SCIENTIFIC REPORTS. 2018;8. doi: 10.1038/s41598-018-25305-y.

**Supplementary Table 5:** Citations of studies excluded from full text screening.

Al-Dabhani K., Tsilidis K., Gunter M., Tzoulaki I. Association between vitamin D and colorectal cancer: A 2-sample Mendelian randomization approach. Mutagenesis. 2018;33(4):e5–6.

Anderson E, Wade KH, Hemani G, Bowden J, Korologou-Linden R, Davey Smith G, et al. The Causal Effect Of Educational Attainment On Alzheimer’s Disease: A Two-Sample Mendelian Randomization Study [Internet]. Epidemiology; 2017. Available from: <http://biorxiv.org/lookup/doi/10.1101/127993>

Bartels M., Baselmans B. First genetic variants for eudaimonia and the genetic overlap with hedonia. Behav Genet. 2018;48(6):456.

Bowden J, Spiller W, Del Greco F, Sheehan N, Thompson J, Minelli C, et al. Improving the visualization, interpretation and analysis of two-sample        summary data Mendelian randomization via the Radial plot and Radial        regression. INTERNATIONAL JOURNAL OF EPIDEMIOLOGY. 2018;47(4):1264–78.

Brumpton B, Sanderson E, Hartwig FP, Harrison S, Vie GÅ, Cho Y, et al. Within-family studies for Mendelian randomization: avoiding dynastic, assortative mating, and population stratification biases [Internet]. Genetics; 2019. Available from: <http://biorxiv.org/lookup/doi/10.1101/602516>

Budu-Aggrey A., Brumpton B., Tyrell J., Watkins S., Frayling T., Asvold B., et al. Investigating a causal relationship between body mass index and inflammatory skin disease using mendelian randomisation. J Invest Dermatol. 2018;138(5):S73.

Budu-Aggrey A, Brumpton B, Tyrrell J, Watkins S, Modalsli E, Celis-Morales C, et al. Evidence of a causal relationship between body mass index and psoriasis:        A mendelian randomization study. PLOS MEDICINE. 2019;16(1).

Burgess S., Davey Smith G. Mendelian Randomization Implicates High-Density Lipoprotein Cholesterol-Associated Mechanisms in Etiology of Age-Related Macular Degeneration. Ophthalmology. 2017;124(8):1165–74.

Byrne EM, Yang J, Wray NR. Inference in Psychiatry via 2-Sample Mendelian Randomization-From Association to Causal Pathway?. JAMA Psychiatry. 2017;74(12):1191–2.

Cao W., Li X., Zhang X., Zhang J., Sun Q., Xu X., et al. No causal effect of telomere length on ischemic stroke and its subtypes: A Mendelian randomization study. Cells. 2019;8(2):159.

Caramaschi D, Taylor AE, Richmond RC, Havdahl KA, Golding J, Relton CL, et al. Maternal smoking during pregnancy and autism: using causal inference methods in a birth cohort study. Translational Psychiatry. 2018;8(1):262.

Carreras-Torres R, Haycock PC, Relton CL, Martin RM, Smith GD, Kraft P, et al. The causal relevance of body mass index in different histological types of lung cancer: A Mendelian randomization study. Sci rep. 2016;6(101563288):31121.

Carreras-Torres R, Johansson M, Haycock PC, Relton CL, Davey Smith G, Brennan P, et al. Role of obesity in smoking behaviour: Mendelian randomisation study in UK Biobank. BMJ. 2018;k1767.

Carter AR, Gill D, Davies NM, Taylor AE, Tillmann T, Vaucher J, et al. What explains the effect of education on cardiovascular disease? Applying Mendelian randomization to identify the consequences of education inequality: [Internet]. Epidemiology; 2018. Available from: <http://biorxiv.org/lookup/doi/10.1101/488254>

Carvalho CM, Wendt FR, Stein DJ, Stein MB, Gelernter J, Belangero SI, et al. Metabolome-Wide Mendelian Randomization Analysis of Emotional and Behavioral Responses to Traumatic Stress. bioRxiv. 2019;545442.

Chen C.-Y., Smoller J., Neale B. Investigating the causal relationships between mental and physical health outcomes using Mendelian randomization. Behav Genet. 2018;48(6):462.

Cheung C.-L., Tan K.C.B., Au P.C.M., Li G.H.Y., Cheung B.M.Y. Evaluation of GDF15 as a therapeutic target of cardiometabolic diseases in human: A Mendelian randomization study. EBioMedicine. 2019;41:85–90.

Cho Y, Haycock PC, Gaunt TR, Zheng J, Morris AP, Davey Smith G, et al. MR-TRYX: Exploiting horizontal pleiotropy to infer novel causal pathways [Internet]. Bioinformatics; 2018. Available from: <http://biorxiv.org/lookup/doi/10.1101/476085>

Corbin L.J., Richmond R.C., Wade K.H., Burgess S., Bowden J., Smith G.D., et al. BMI as a modifiable risk factor for type 2 diabetes: Refining and understanding causal estimates using mendelian randomization. Diabetes. 2016;65(10):3002–7.

Crawford A.A., Walker B.R. Identifying and utilising genetic variants associated with morning plasma cortisol: A cortisol network (CORNET) analysis. Genet Epidemiol. 2017;41(7):668–9.

Dardani C, Howe L, Stergiakouli E, Wren Y, Humphries K, Davies A, et al. Cleft lip/palate and educational attainment: cause, consequence, or correlation? A Mendelian randomization study: [Internet]. Genetics; 2018. Available from: <http://biorxiv.org/lookup/doi/10.1101/434126>

Ding R, Huang T, Han J, Ding R, Huang T, Han J. Diet/lifestyle and risk of diabetes and glycemic traits: a Mendelian        randomization study. LIPIDS IN HEALTH AND DISEASE. 2018;17.

Disney-Hogg L, Cornish AJ, Sud A, Law PJ, Kinnersley B, Jacobs DI, et al. Impact of atopy on risk of glioma: a Mendelian randomisation study. BMC Med. 2018;16(1):42.

Dixon P, Hollingworth W, Harrison S, Davies NM, Smith GD. The causal effect of adiposity on hospital costs: Mendelian Randomization analysis of over 300,000 individuals from the UK Biobank [Internet]. Epidemiology; 2019. Available from: <http://biorxiv.org/lookup/doi/10.1101/589820>

Dudding T., Johansson M., Thomas S.J., Brennan P., Martin R.M., Timpson N.J. Assessing the causal association between 25-hydroxyvitamin D and the risk of oral and oropharyngeal cancer using Mendelian randomization. Int J Cancer. 2018;143(5):1029–36.

Ellervik C, Roselli C, Christophersen I, Alonso A, Pietzner M, Sitlani C, et al. Assessment of the Relationship Between Genetic Determinants of Thyroid        Function and Atrial Fibrillation A Mendelian Randomization Study. JAMA CARDIOLOGY. 2019;4(2):144–52.

Emanuelsson F., Nordestgaard B., Tybjaerg-Hansen A., Benn M. High LDL cholesterol levels and risk of peripheral vascular diseases: A Mendelian randomisation study including 116,419 individuals from the general population. Diabetologia. 2018;61:S136–7.

Evans DM, Moen G-H, Hwang L-D, Lawlor DA, Warrington NM. Elucidating the role of maternal environmental exposures on offspring health and disease using two-sample Mendelian randomization. International Journal of Epidemiology [Internet]. 2019; Available from: <https://academic.oup.com/ije/advance-article/doi/10.1093/ije/dyz019/5366230>

Fang X, Zhou J, Cai J, Che C, Xaing E, Li H, et al. Childhood Obesity Leads to Adult Type 2 Diabetes and Coronary Artery Diseases: A Two-Sample Mendelian Randomization Study [Internet]. Rochester, NY: Social Science Research Network; 2018. Report No.: ID 3247843. Available from: <https://papers.ssrn.com/abstract=3247843>

Fatima T. Gout and Metabolic Disease: Investigation of Potential Relationship in the New Zealand Population [Internet]. University of Otago; 2017. Available from: <https://ourarchive.otago.ac.nz/handle/10523/7708>

Fluharty ME, Sallis H, Munafo MR, Aklin A-M Bagot, Bowden, Brazil, Breyer, Brody, Brook, Brook, Burgess, Burgess, Burgess, Cadoret, Corbin, Corrigall, Crone, Dalley, Davey Smith, Demontis, Dishion, Disney, Dresel, Farrell, Gage, Gage, Gage, Gehricke, Goldstein, Grace, Hicks, Holmes, John, Kim, Kliewer, Kollins, Korhonen, Krause, Krause, Krause, Kretschmer, Lambert, Lasser, Laucht, Lawlor, Levin, Mcardle, Mcclernon, Mcclernon, Mick, Milberger, Mostafavi, Munafo, Pappa, Pomerleau, Potter, Rodriguez, Rohde, Solanto, Spoth, Steinberg, Sterne, Verdejo-Garcia, Volkow, Volkow, Wang. Investigating possible causal effects of externalizing behaviors on tobacco initiation: A Mendelian randomization analysis. Drug and Alcohol Dependence. 2018;191:338–42.

Gage S., Jones H., Taylor A., Zammit S., Munafo M. Using publicly available data to investigate causal associations between substance use and mental health. Eur Neuropsychopharmacol. 2016;26:S57–8.

Gage S.H., Jones H.J., Burgess S., Bowden J., Davey Smith G., Zammit S., et al. Assessing causality in associations between cannabis use and schizophrenia risk: a two-sample Mendelian randomization study. Psychol Med. 2017;47(5):971–80.

Gage SH, Bowden J, Davey Smith G, Munafo MR. Investigating causality in associations between education and smoking: a two-sample Mendelian randomization study. Int J Epidemiol. 2018;47(4):1131–40.

Georgakis M, Gill D, Rannikmae K, Traylor M, Anderson C, Lee J, et al. Genetically Determined Levels of Circulating Cytokines and Risk of        Stroke: Role of Monocyte Chemoattractant Protein-1. CIRCULATION. 2019;139(2):256–68.

Gill D, Del Greco F, Walker A, Srai S, Laffan M, Minelli C, et al. The Effect of Iron Status on Risk of Coronary Artery Disease A Mendelian        Randomization Study-Brief Report. ARTERIOSCLEROSIS THROMBOSIS AND VASCULAR BIOLOGY. 2017;37(9):1788-+.

Gill D, Georgakis M, Laffan M, Sabater-Lleal M, Malik R, Tzoulaki I, et al. Genetically Determined FXI (Factor XI) Levels and Risk of Stroke. STROKE. 2018;49(11):2761–3.

Gill D. Blood pressure traits differentially affect risk of different ischaemic stroke subtypes. Eur Stroke J. 2018;3(1):58–9.

Gill D., Brewer C.F., Del Greco M F., Sivakumaran P., Bowden J., Sheehan N.A., et al. Age at menarche and adult body mass index: a Mendelian randomization study. Int J Obes. 2018;42(9):1574–81.

Goto A, Yamaji T, Sawada N, Momozawa Y, Kamatani Y, Kubo M, et al. Diabetes and Cancer Risk: A Mendelian Randomization Study. International Journal of Cancer [Internet]. 2019 [cited 4AD Jan 1]; Available from: <https://onlinelibrary.wiley.com/doi/abs/10.1002/ijc.32310>

Grover S., Del Greco M.F., Lill C., Konig I.R. Modifiable risk factors and Parkinson’s disease: systematic Mendelian randomization studies. Genet Epidemiol. 2018;42(7):703.

Gupta V, Sachdeva MP, Walia GK. “Mendelian Randomization” Approach in Economic Assessment of Health Conditions. Front Public Health [Internet]. 2019;7. Available from: <https://www.ncbi.nlm.nih.gov/pmc/articles/PMC6369183/>

Hagenaars S, Gale C, Deary I, Harris S, Hagenaars SP, Gale CR, et al. Cognitive ability and physical health: a Mendelian randomization study. SCIENTIFIC REPORTS. 2017;7.

Harati H., Zanetti D., Ingelsson E., Knowles J. No evidence of a causal association between diabetes-related phenotypes and atrial fibrillation. Diabetes. 2018;67:A460.

Harati H, Zanetti D, Rao A, Gustafsson S, Perez M, Ingelsson E, et al. No evidence of a causal association of type 2 diabetes and glucose metabolism with atrial fibrillation. Diabetologia. 2019;62(5):800–4.

Harshfield E.L., Stacey D., Paul D.S., Koulman A., Wood A.M., Butterworth A.S., et al. Genomics of lipid metabolism: Identifying novel causal pathways and new therapeutic targets for reducing risk of coronary heart disease. Genet Epidemiol. 2016;40(7):614–5.

Hartwig F.P., Steinberg J., Zengini E., Hatzikotoulas K., Southam L., Tachmazidou I., et al. Causal effects for higher body mass index, but not for triglyceride levels or genetic predisposition to type 2 diabetes, on osteoarthritis. Osteoarthritis Cartilage. 2018;26:S29–30.

Hartwig F, Borges M, Horta B, Bowden J, Smith G, Hartwig FP, et al. Inflammatory Biomarkers and Risk of Schizophrenia A 2-Sample Mendelian        Randomization Study. JAMA PSYCHIATRY. 2017;74(12):1226–33.

Hartwig F, Bowden J, de Mola C, Tovo-Rodrigues L, Smith G, Horta B, et al. Body mass index and psychiatric disorders: a Mendelian randomization        study. SCIENTIFIC REPORTS. 2016;6.

Hartwig F, Davies N, Hemani G, Smith G, Hartwig FP, Davies NM, et al. Two-sample Mendelian randomization: avoiding the downsides of a        powerful, widely applicable but potentially fallible technique. INTERNATIONAL JOURNAL OF EPIDEMIOLOGY. 2016;45(6):1717–26.

Haworth S., Haycock P., West N., Thomas S., Franks P., Timpson N. Gene discovery for oral ulceration: A UK Biobank Study. Lancet. 2017;389:S46.

Haycock PC, Burgess S, Nounu A, Zheng J, Okoli GN, Bowden J, et al. Association Between Telomere Length and Risk of Cancer and Non-Neoplastic Diseases: A Mendelian Randomization Study. JAMA Oncol. 2017;3(5):636–51.

Hemani G, Bowden J, Haycock PC, Zheng J, Davis O, Flach P, et al. Automating Mendelian randomization through machine learning to construct a putative causal map of the human phenome [Internet]. Epidemiology; 2017. Available from: <http://biorxiv.org/lookup/doi/10.1101/173682>

Hillary RF, McCartney DL, Harris SE, Stevenson AJ, Seeboth A, Zhang Q, et al. Genetic and epigenetic architectures of neurological protein biomarkers in the Lothian Birth Cohort 1936. bioRxiv. 2019;558940.

Holmes MV, Davey Smith G. Challenges in Interpreting Multivariable Mendelian Randomization: Might “Good Cholesterol” Be Good After All? American Journal of Kidney Diseases. 2018;71(2):149–53.

Howe LJ, Richardson TG, Arathimos R, Alvizi L, Passos-Bueno MR, Stanier P, et al. Evidence for DNA methylation mediating genetic liability to non-syndromic cleft lip/palate. Epigenomics. 2019;11(2):133–45.

Howe LJ, Lawson DJ, Davies NM, Pourcain BS, Lewis SJ, Smith GD, et al. Alcohol consumption and mate choice in UK Biobank: comparing observational and Mendelian randomization estimates. bioRxiv. 2019;418269.

Howell AE, Zheng J, Haycock PC, McAleenan A, Relton C, Martin RM, et al. Use of Mendelian Randomization for Identifying Risk Factors for Brain Tumors. Front Genet [Internet]. 2018;9. Available from: <https://www.ncbi.nlm.nih.gov/pmc/articles/PMC6240585/>

Hu X, Zhuang X, Mei W, Liu G, Du Z, Liao X, et al. Exploring the Causal Pathway From Body Mass Index to Coronary Heart Disease: A Network Mendelian Randomization Study [Internet]. Rochester, NY: Social Science Research Network; 2019. Report No.: ID 3343625. Available from: <https://papers.ssrn.com/abstract=3343625>

Huang J, Schooling C, Huang JV, Schooling CM. Inflammation and bone mineral density: A Mendelian randomization study. SCIENTIFIC REPORTS. 2017;7.

Huang T., Geng T. Maternal central obesity and birth size-a mendelian randomization analysis. Diabetes. 2018;67:A383.

Hypponen E, Gebremichael AM, Zhou A, Santhanakrishnan VK. Phenome-Wide Analysis to Establish Causal Evidence for an Association Between Body Mass Index and Multiple Disease Outcomes in the UK Biobank [Internet]. Rochester, NY: Social Science Research Network; 2019. Report No.: ID 3354708. Available from: <https://papers.ssrn.com/abstract=3354708>

Jones H., Smith G.D., O’Donovan M.C., Owen M.J., Walters J., Zammit S. Investigating a causal association between neuroticism and schizophrenia using two-sample mendelian randomization. Schizophr Bull. 2018;44:S273–4.

Kar SP, Brenner H, Giles GG, Huo D, Milne RL, Rennert G, et al. The association between low-density lipoprotein cholesterol predicted by HMGCR genetic variants and breast cancer risk may be mediated by body mass index. bioRxiv. 2018;464446.

Kia DA, Noyce AJ, White J, Speed D, Nicolas A, IPDGC collaborators, et al. Mendelian randomization study shows no causal relationship between circulating urate levels and Parkinson’s disease. Ann Neurol. 2018;84(2):191–9.

Kwok M, Leung G, Schooling C, Kwok MK, Leung GM, Schooling CM. Habitual coffee consumption and risk of type 2 diabetes, ischemic heart        disease, depression and Alzheimer’s disease: a Mendelian randomization        study. SCIENTIFIC REPORTS. 2016;6.

Lai F, Nath M, Hamby S, Thompson J, Nelson C, Samani N, et al. Adult height and risk of 50 diseases: a combined epidemiological and        genetic analysis. BMC MEDICINE. 2018;16.

Lanktree M, Theriault S, Walsh M, Pare G, Lanktree MB, Theriault S, et al. HDL Cholesterol, LDL Cholesterol, and Triglycerides as Risk Factors for        CKD: A Mendelian Randomization Study. AMERICAN JOURNAL OF KIDNEY DISEASES. 2018;71(2):166–72.

Larsson S, Markus H, Larsson SC, Markus HS. Branched-chain amino acids and Alzheimer’s disease: a Mendelian        randomization analysis. SCIENTIFIC REPORTS. 2017;7.

Lee YS, Cho Y, Burgess S, Davey Smith G, Relton CL, Shin S-Y, et al. Serum gamma-glutamyl transferase and risk of type 2 diabetes in the general Korean population: a Mendelian randomization study. Hum Mol Genet. 2016;25(17):3877–86.

Li S., Teumer A., Knol M.J., Yang Q., Seshadri S. GENOME-WIDE ASSOCIATION STUDY OF 11,785 INDIVIDUALS IDENTIFIES SEVEN LOCI ASSOCIATED WITH BRAIN-DERIVED NEUROTROPHIC FACTOR. Alzheimer’s Dementia. 2018;14(7):P262.

Li S. Mendelian Randomization when Many Instruments are Invalid: Hierarchical Empirical Bayes Estimation. arXiv:170601389 [stat] [Internet]. 2017; Available from: <http://arxiv.org/abs/1706.01389>

Liu J., Zhou H., Zhang Y., Huang Y., Fang W., Yang Y., et al. Docosapentaenoic Acid and Lung Cancer Risk: A Mendelian Randomisation Study. J Thorac Oncol. 2018;13(12):S1065.

Llano A., Tan L.E., Warren H., McCallum L., Dominiczak A.F., Padmanabhan S. Investigating causal relationships of blood pressure on type 2 diabetes using mendelian randomisation. J Hypertens. 2018;36:e258.

lloyd. Bidirectional effects of anxiety and anorexia nervosa: A Mendelian randomization study. 2018;

Mack S, Coassin S, Vaucher J, Kronenberg F, Lamina C, Mack S, et al. Evaluating the Causal Relation of ApoA-IV with Disease-Related Traits -        A Bidirectional Two-sample Mendelian Randomization Study. SCIENTIFIC REPORTS. 2017;7.

Magnus M.C., Miliku K., Bauer A., Engel S.M., Felix J.F., Jaddoe V.W.V., et al. Vitamin D and Risk of Pregnancy-Related Hypertensive Disorders: Mendelian Randomization Study. Obstet Gynecol Surv. 2018;73(11):617–9.

Magnus M, Miliku K, Bauer A, Engel S, Felix J, Jaddoe V, et al. Vitamin D and risk of pregnancy related hypertensive disorders:        mendelian randomisation study. BMJ-BRITISH MEDICAL JOURNAL. 2018;361.

McGowan LM, Davey Smith G, Gaunt TR, Richardson TG. Integrating Mendelian randomization and multiple-trait colocalization to uncover cell-specific inflammatory drivers of autoimmune and atopic disease [Internet]. Genetics; 2018. Available from: <http://biorxiv.org/lookup/doi/10.1101/394296>

Meyer HV, Dawes TJ, Serrani M, Bai W, Tokarczuk P, Cai J, et al. Genomic analysis reveals a functional role for myocardial trabeculae in adults [Internet]. Genomics; 2019. Available from: <http://biorxiv.org/lookup/doi/10.1101/553651>

Milaneschi Y, Peyrot WJ, Nivard MG, Mbarek H, Boomsma DI, Penninx BW. A role for vitamin D and omega-3 fatty acids in major depression? An exploration using genomics. bioRxiv. 2019;516013.

Mohammadi-Shemirani P, Sjaarda J, Gerstein H, Treleaven D, Walsh M, Mann J, et al. A Mendelian Randomization-Based Approach to Identify Early and Sensitive        Diagnostic Biomarkers of Disease. CLINICAL CHEMISTRY. 2019;65(3):427–36.

Mokry L, Ross S, Timpson N, Sawcer S, Smith G, Richards J, et al. Obesity and Multiple Sclerosis: A Mendelian Randomization Study. PLOS MEDICINE. 2016;13(6).

Morales E, Vilahur N, Salas LA, Motta V, Fernandez MF, Murcia M, et al. Genome-wide DNA methylation study in human placenta identifies novel loci associated with maternal smoking during pregnancy. Int J Epidemiol. 2016;45(5):1644–55.

Morris AP, Le TH, Wu H, Akbarov A, Most PJ van der, Hemani G, et al. Trans-ethnic genome-wide association study of kidney function provides novel insight into effector genes and causal effects on kidney-specific disease aetiologies. bioRxiv. 2018;420273.

Noordam R., Oudt C.H., Bos M.M., Smit R.A.J., van Heemst D. High-sensitivity C-reactive protein, low-grade systemic inflammation and type 2 diabetes mellitus: A two-sample Mendelian randomization study. Nutr Metab Cardiovasc Dis. 2018;28(8):795–802.

Noordam R, Smit RA, Postmus I, Trompet S, van Heemst D. Lessons from Mendelian randomization studies on liver biomarkers: response to Abbasi. International Journal of Epidemiology. 2017;46(5):1713–4.

Noyce A., Kia D., Heilbron K., Jepson J., Hemani G., Hinds D., et al. Tendency towards being a “Morning person” increases risk of Parkinson’s disease: Evidence from Mendelian randomisation. Mov Disord. 2018;33:S364.

Noyce A., Kia D., Hemani G., Nicolas A., Price T., Fernandez E., et al. Increased BMI may protect against parkinson’s disease: Evidence from mendelian randomisation study. Mov Disord. 2017;32:5.

Noyce A, Kia D, Hemani G, Nicolas A, Price T, De Pablo-Fernandez E, et al. Estimating the causal influence of body mass index on risk of Parkinson        disease: A Mendelian randomisation study. PLOS MEDICINE. 2017;14(6).

Noyce AJ, Bandres-Ciga S, Kim J, Heilbron K, Kia D, Hemani G, et al. The Parkinson’s Disease Mendelian Randomization Research Portal. bioRxiv. 2019;604033.

O’Connor LJ, Price AL. Distinguishing genetic correlation from causation across 52 diseases and complex traits [Internet]. Genetics; 2017. Available from: <http://biorxiv.org/lookup/doi/10.1101/205435>

Ogawa K, Stuart PE, Tsoi LC, Suzuki K, Nair RP, Mochizuki H, et al. A Transethnic Mendelian Randomization Study Identifies Causality of Obesity on Risk of Psoriasis. Journal of Investigative Dermatology [Internet]. 2018; Available from: <http://www.sciencedirect.com/science/article/pii/S0022202X18329154>

Ong J, Hwang L, Cuellar-Partida G, Martin N, Chenevix-Trench G, Quinn M, et al. Assessment of moderate coffee consumption and risk of epithelial ovarian        cancer: a Mendelian randomization study. INTERNATIONAL JOURNAL OF EPIDEMIOLOGY. 2018;47(2):450–9.

Parisinos C., Serghiou S., Katsoulis M., Patel R., Hemingway H., Hingorani A. The interleukin-6 receptor as a drug target in inflammatory bowel disease; a mendelian randomization study. United Eur Gastroenterol J. 2018;6(8):A42–3.

Parisinos C.A., Serghiou S., Katsoulis M., Patel R.S., Hemingway H., Hingorani A.D. The interleukin-6 receptor as a target for prevention of Crohn’s disease and ulcerative colitis; A Mendelian randomisation study. J Crohn’s Colitis. 2018;12:S530–1.

Pasman JA, Verweij KJH, Gerring Z, Stringer S, Sanchez-Roige S, Treur JL, et al. Genome-wide association analysis of lifetime cannabis use (N=184,765) identifies new risk loci, genetic overlap with mental health, and a causal influence of schizophrenia on cannabis use [Internet]. Genetics; 2018. Available from: <http://biorxiv.org/lookup/doi/10.1101/234294>

Polimanti R, Ratanatharathorn A, Maihofer AX, Choi KW, Stein MB, Morey RA, et al. Economic status mediates the relationship between educational attainment and posttraumatic stress disorder: a multivariable Mendelian randomization study: Supplemental Material [Internet]. Genetics; 2018. Available from: <http://biorxiv.org/lookup/doi/10.1101/503300>

Porcu E, Rüeger S, Lepik K, Consortium eQTLGen, Santoni FA, Reymond A, et al. Mendelian Randomization integrating GWAS and eQTL data reveals genetic determinants of complex and clinical traits. bioRxiv. 2019;377267.

PRACTICAL consortium, Yarmolinsky J, Berryman K, Langdon R, Bonilla C, Davey Smith G, et al. Mendelian randomization does not support serum calcium in prostate cancer risk. Cancer Causes & Control. 2018;29(11):1073–80.

Prats-Uribe A., Sayols-Baixeras S., Fernandez-Sanles A., Duarte-Salles T., Logue J., Elosua R., et al. The causal association between childhood and adulthood body mass index and osteoarthritis: A mendelian randomization study. Ann Rheum Dis. 2018;77:1188–9.

Ravera S., Carrasco N., Gelernter J., Polimanti R. A Mendelian randomization phenome-wide association study of thyroid function in 337,199 individuals from UK Biobank. Hum Genomics. 2018;12.

Richardson TG, Harrison S, Hemani G, Smith GD. An atlas of polygenic risk score associations to highlight putative causal relationships across the human phenome. :24.

Richardson TG, Haycock PC, Zheng J, Timpson NJ, Gaunt TR, Davey Smith G, et al. Systematic Mendelian randomization framework elucidates hundreds of genetic loci which may influence disease through changes in DNA methylation levels [Internet]. Genetics; 2017. Available from: <http://biorxiv.org/lookup/doi/10.1101/189076>

Richardson TG, Zheng J, Davey Smith G, Timpson NJ, Gaunt TR, Relton CL, et al. Causal epigenome-wide association study identifies CpG sites that influence cardiovascular disease risk [Internet]. Epidemiology; 2017. Available from: <http://biorxiv.org/lookup/doi/10.1101/132019>

Richardson TG, Harrison S, Hemani G, Smith GD. An atlas of polygenic risk score associations to highlight putative causal relationships across the human phenome. :24.

Richmond R., Anderson E., Dashti H., Jones S., Lane J., Relton C., et al. Investigating causal relationships between sleep characteristics and risk of breast cancer: A Mendelian randomization study. Br J Cancer. 2018;119(1):35.

Richmond RC, Anderson EL, Dashti HS, Jones SE, Lane JM, Strand LB, et al. Investigating causal relationships between sleep traits and risk of breast cancer: a Mendelian randomization study [Internet]. Epidemiology; 2018. Available from: <http://biorxiv.org/lookup/doi/10.1101/457572>

Richmond R, Wade K, Corbin L, Bowden J, Hemani G, Timpson N, et al. Investigating the role of insulin in increased adiposity: Bi-directional Mendelian randomization study [Internet]. Epidemiology; 2017. Available from: <http://biorxiv.org/lookup/doi/10.1101/155739>

Rosoff DB, Lohoff F. Educational attainment causally impacts drinking behaviors and risk for alcohol dependence: results from a two-sample Mendelian randomization study in ~ 780,000 study participants [Internet]. Epidemiology; 2019. Available from: <http://biorxiv.org/lookup/doi/10.1101/557595>

Sabater-Lleal M., Huffman J.E., de Vries P.S., Marten J., Mastrangelo M.A., Song C., et al. Genome-Wide Association Transethnic Meta-Analyses Identifies Novel Associations Regulating Coagulation Factor VIII and von Willebrand Factor Plasma Levels. Circulation. 2019;139(5):620–35.

Sallis H., Davey Smith G., Munafo M. SMOKING AND NEUROTICISM: USING MENDELIAN RANDOMIZATION TO INVESTIGATE CAUSALITY. Eur Neuropsychopharmacol. 2019;29:S1018–9.

Sandu M.R., Beynon R., Richmond R., Santos Ferreira D.L., Athene Lane J., Martin R.M. A novel application of two-step Mendelian randomization: Applying the results of small feasibility studies of interventions to infer causal effects on clinical endpoints. Br J Cancer. 2018;119(1):34–5.

Schooling C, Ng J. Reproduction and longevity A Mendelian randomization study of gonadotropin-releasing hormone and ischemic heart disease [Internet]. Evolutionary Biology; 2018. Available from: <http://biorxiv.org/lookup/doi/10.1101/472548>

So H-C, Cheng Y, Chau CK, Sham PC. Causal relationships between blood lipids and depression phenotypes: A Mendelian randomization analysis: Supplementary Tables [Internet]. Genetics; 2018. Available from: <http://biorxiv.org/lookup/doi/10.1101/363119>

Song GG, Lee YH. Causal Association between Bone Mineral Density and Osteoarthritis: A Mendelian Randomization Study. Journal of Rheumatic Diseases. 2019;26(2):104–10.

Speed D, Hemani G, Speed MS, Major Depressive Disorder Working Group of the Psychiatric Genomics Consortium, Boerglum AD, Oestergaard SD. Does Neuroticism Cause Depression? A Mendelian Randomization Study [Internet]. Genetics; 2018. Available from: <http://biorxiv.org/lookup/doi/10.1101/420703>

Sun D., Richard M., Ding J., Launer L.J., Sidney S., Yaffe K., et al. INVESTIGATING THE CAUSAL ASSOCIATION OF BLOOD PRESSURE AND MIDLIFE COGNITIVE FUNCTION USING MENDELIAN RANDOMIZATION. Alzheimer’s Dementia. 2018;14(7):P1110–1.

Sun Y-Q, Brumpton BM, Bonilla C, Lewis SJ, Burgess S, Skorpen F, et al. Serum 25-hydroxyvitamin D levels and risk of lung cancer and histologic types: a Mendelian randomisation analysis of the HUNT study. European Respiratory Journal. 2018;51(6):1800329.

Takahashi H, Cornish A, Sud A, Law P, Kinnersley B, Ostrom Q, et al. Mendelian randomisation study of the relationship between vitamin D and        risk of glioma. SCIENTIFIC REPORTS. 2018;8.

Takahashi H, Cornish AJ, Sud A, Law PJ, Disney-Hogg L, Calvocoressi L, et al. Mendelian randomization provides support for obesity as a risk factor for meningioma. Sci Rep [Internet]. 2019;9. Available from: <https://www.ncbi.nlm.nih.gov/pmc/articles/PMC6343031/>

Tan L.E., Aman A., McCallum L., Dominiczak A.F., Padmanabhan S. Causal association of uromodulin with blood pressure independent of renal function. J Hypertens. 2018;36:e345–6.

Tan L.E., Llano A., Aman A., Dominiczak A.F., Padmanabhan S. Mendelian randomization study of causal relationship of height on blood pressure and arterial stiffness. J Hypertens. 2018;36:e91–2.

Teumer A, Gambaro G, Corre T, Bochud M, Vollenweider P, Guessous I, et al. Negative effect of vitamin D on kidney function: a Mendelian        randomization study. NEPHROLOGY DIALYSIS TRANSPLANTATION. 2018;33(12):2139–45.

Timms J.A., Relton C.L., Rankin J., Strathdee G., McKay J.A. Environment, DNA methylation and risk of childhood acute lymphoblastic leukaemia: a novel Mendelian randomization study. Mutagenesis. 2018;33(4):e7.

Trajanoska K, Morris JA, Oei L, Zheng H-F, Evans DM, Kiel DP, et al. Assessment of the genetic and clinical determinants of fracture risk: genome wide association and mendelian randomisation study. BMJ. 2018;362(8900488):k3225.

Valdes-Marquez E., Parish S., Clarke R., Stari T., Worrall B.B., Hopewell J.C. Relative effects of LDL-C on ischemic stroke and coronary disease: A Mendelian randomization study. Neurology. 2019;92(11):e1176–87.

van Vliet N, Noordam R, van Klinken J, Westendorp R, Bassett J, Williams G, et al. Thyroid Stimulating Hormone and Bone Mineral Density: Evidence From a        Two-Sample Mendelian Randomization Study and a Candidate Gene        Association Study. JOURNAL OF BONE AND MINERAL RESEARCH. 2018;33(7):1318–25.

Verweij KJH, Treur JL, Vink JM. Investigating causal associations between use of nicotine, alcohol, caffeine and cannabis: a two-sample bidirectional Mendelian randomization study. Addiction. 2018;113(7):1333–8.

Walker V.M., Davies N.M., Kehoe P.G., Martin R.M. Can treatments for hypertension be repurposed for the treatment of dementia? Pharmacoepidemiol Drug Saf. 2018;27:194.

Walker VM, Kehoe PG, Martin RM, Davies NM. Repurposing antihypertensive drugs for the prevention of Alzheimer’s disease: a Mendelian Randomization study [Internet]. Genetics; 2018. Available from: <http://biorxiv.org/lookup/doi/10.1101/486878>

Wang H, Lane JM, Jones SE, Dashti HS, Ollila H, Wood AR, et al. Genome-wide association analysis of excessive daytime sleepiness identifies 42 loci that suggest phenotypic subgroups [Internet]. Genomics; 2018. Available from: <http://biorxiv.org/lookup/doi/10.1101/454561>

Wang J, Kwok MK, Au Yeung SL, Li AM, Lam HS, Leung JYY, et al. Sleep duration and risk of diabetes: Observational and Mendelian randomization studies. Preventive Medicine. 2019;119:24–30.

Ward-Caviness CK, de Vries PS, Wiggins KL, Huffman JE, Yanek LR, Bielak LF, et al. Evaluation of the causal effect of fibrinogen on incident coronary heart disease via Mendelian randomization [Internet]. Genetics; 2018. Available from: <http://biorxiv.org/lookup/doi/10.1101/448381>

Ware JJ, Tanner J-A, Taylor AE, Bin Z, Haycock P, Bowden J, et al. Does coffee consumption impact on heaviness of smoking? Addiction. 2017;112(10):1842–

Wendt FR, Carvalho C, Gelernter J, Polimanti R. DRD2 and FOXP2 are implicated in the associations between computerized device use and psychiatric disorders [Internet]. Genomics; 2018. Available from: <http://biorxiv.org/lookup/doi/10.1101/497420>

Weng L, Roetker N, Lutsey P, Alonso A, Guan W, Pankow J, et al. Evaluation of the relationship between plasma lipids and abdominal        aortic aneurysm: A Mendelian randomization study. PLOS ONE. 2018;13(4).

Wiklund P, Karhunen V, Richmond R, Rodriguez A, De Silva M, Wielscher M, et al. DNA methylation links prenatal smoking exposure to later life health outcomes in offspring. [Internet]. Epidemiology; 2018. Available from: <http://biorxiv.org/lookup/doi/10.1101/428896>

Wood A., Guggenheim J.A. Refractive error has minimal influence on the risk of age-related macular degeneration: A Mendelian randomization study. Am J Ophthalmol. 2019;

Wu S, Wang P, Xiao C, Li Z, Yang B, Fu J, et al. A Quick-responsive DNA Nanotechnology Device for Bio-molecular        Homeostasis Regulation. SCIENTIFIC REPORTS. 2016;6.

Xu L, Lin S, Schooling C, Xu L, Lin SL, Schooling CM. A Mendelian randomization study of the effect of calcium on coronary        artery disease, myocardial infarction and their risk factors. SCIENTIFIC REPORTS. 2017;7.

Xu L., Borges M.C., Hemani G., Lawlor D.A. The role of glycaemic and lipid risk factors in mediating the effect of BMI on coronary heart disease: a two-step, two-sample Mendelian randomisation study. Diabetologia. 2017;60(11):2210–20.

Yarmolinsky J, Relton CL, Lophatananon A, Muir K, Menon U, Gentry-Maharaj A, et al. Evaluating causal associations between previously reported risk factors and epithelial ovarian cancer: a Mendelian randomization analysis [Internet]. Epidemiology; 2018. Available from: <http://biorxiv.org/lookup/doi/10.1101/472696>

Yin P, Anttila V, Siewert KM, Palotie A, Davey Smith G, Voight BF. Serum calcium and risk of migraine: a Mendelian randomization study. Hum Mol Genet. 2017;26(4):820–8.

zen. Causal Association between Birth Weight and Adult Diseases: Evidence from a Mendelian Randomisation Analysis. 2018;

Zhan Y., Karlsson I.K., Karlsson R., Tillander A., Reynolds C.A., Pedersen N.L., et al. Exploring the Causal Pathway from Telomere Length to Coronary Heart Disease: A Network Mendelian Randomization Study. Circ Res. 2017;121(3):214–9.

Zhao Q, Wang J, Hemani G, Bowden J, Small DS. Statistical inference in two-sample summary-data Mendelian randomization using robust adjusted profile score. arXiv:180109652 [math, stat] [Internet]. 2018; Available from: <http://arxiv.org/abs/1801.09652>

Zheng J., Brion M.-J., Kemp J., Warrington N., Hemani G., Qiao Z., et al. Do blood lipid levels influence bone mineral density? Findings from a Mendelian randomization study. J Musculoskelet Neuronal Interact. 2018;18(1):127–8.

Zheng J., Haycock P., Hemani G., Elsworth B., Shihab H., Laurin C., et al. LD hub and MR-base: Online platforms for preforming LD score regression and Mendelian randomization analysis using GWAS summary data. Behav Genet. 2016;46(6):815.

Zhuang H, Han J, Cheng L, Liu S, Zhuang H, Han J, et al. A Positive Causal Influence of IL-18 Levels on the Risk of T2DM: A        Mendelian Randomization Study. FRONTIERS IN GENETICS. 2019;10.

T Richardson, J Zheng, GD Smith, NJ Timpson, TR Gaunt, CL Relton, G Hemani Mendelian Randomization Analysis Identifies CpG Sites as Putative Mediators for Genetic Influences on Cardiovascular Disease Risk | Elsevier Enhanced Reader. 2019 [cited 4AD Jan 1]; Available from: <https://reader.elsevier.com/reader/sd/pii/S0002929717303701?token=1904A3DA6D7DDED2268E848611BFD156C05233494852318F92BEC721EC1BA609E9F3A42F6E473F8F021E29F37573DD63>

The full results of included studies can be found in **Supplementary Table 6**.

**Supplementary Table 7:** List of studies classified as having ten or more phenotypes.

Adams CD, Richmond R, Ferreira DLS, Spiller W, Tan V, Zheng J, et al. Circulating Metabolic Biomarkers of Screen-Detected Prostate Cancer in the ProtecT Study. Cancer Epidemiol Biomarkers Prev. 2019;28(1):208 doi: 10.1158/1055-9965.EPI-18-0079.

Bandres-Ciga S, Noyce AJ, Hemani G, Nicolas A, Calvo A, Mora G, et al. Shared polygenic risk and causal inferences in amyotrophic lateral sclerosis. Ann Neurol. 2019;85(4):470–81.  DOI: [10.1002/ana.25431](https://doi.org/10.1002/ana.25431)

Brumpton BM, Fritsche LG, Zheng J, Nielsen JB, Mannila M, Surakka I, et al. Variation in Serum PCSK9 (Proprotein Convertase Subtilisin/Kexin Type 9), Cardiovascular Disease Risk, and an Investigation of Potential Unanticipated Effects of PCSK9 Inhibition: A Genome-Wide Association Study and Mendelian Randomization Study in the HUNT, Norway. Circulation: Genomic and Precision Medicine [Internet]. 2019;12(1). Available from: <https://www.ahajournals.org/doi/10.1161/CIRCGEN.118.002335>

Carreras-Torres R, Johansson M, Haycock PC, Wade KH, Relton CL, Martin RM, et al. Obesity, metabolic factors and risk of different histological types of lung cancer: A Mendelian randomization study. PLOS ONE. 2017;12(6):e0177875. <https://doi.org/10.1371/journal.pone.0177875>

Dashti H.S., Redline S., Saxena R. Polygenic risk score identifies associations between sleep duration and diseases determined from an electronic medical record biobank. Sleep. 2019;42(3):zsy247. doi: 10.1093/sleep/zsy247.

Hemani G, Tilling K, Davey Smith G. Orienting The Causal Relationship Between Imprecisely Measured Traits Using Genetic Instruments [Internet]. Systems Biology; 2017. Available from: <http://biorxiv.org/lookup/doi/10.1101/117101>

Joshi P, Pirastu N, Kentistou K, Fischer K, Hofer E, Schraut K, et al. Genome-wide meta-analysis associates HLA-DQA1/DRB1 and LPA and lifestyle        factors with human longevity. NATURE COMMUNICATIONS. 2017;8.  doi: 10.1038/s41467-017-00934-5

Morris AP, Le TH, Wu H, Akbarov A, van der Most PJ, Hemani G, et al. Trans-ethnic kidney function association study reveals putative causal genes and effects on kidney-specific disease aetiologies. Nature Communications. 2019;10(1):29. doi: 10.1038/s41467-018-07867-7.

Ravera S., Carrasco N., Gelernter J., Polimanti R. Phenomic impact of genetically-determined euthyroid function and molecular differences between thyroid disorders. J Clin Med. 2018;7(10):296.  <https://doi.org/10.3390/jcm7100296>

Richardson T.G., Haycock P.C., Zheng J., Timpson N.J., Gaunt T.R., Smith G.D., et al. Systematic Mendelian randomization framework elucidates hundreds of CpG sites which may mediate the influence of genetic variants on disease. Hum Mol Genet. 2018;27(18):3293–304. doi: 10.1093/hmg/ddy210.

Shadrina AS, Sharapov SZ, Shashkova TI, Tsepilov YA. Varicose veins of lower extremities: insights from the first large-scale genetic study [Internet]. Genomics; 2018. Available from: <http://biorxiv.org/lookup/doi/10.1101/368365>

Wootton RE, Lawn RB, Millard LAC, Davies NM, Taylor AE, Munafò MR, et al. Evaluation of the causal effects between subjective wellbeing and cardiometabolic health: mendelian randomisation study. BMJ. 2018;362:k3788. doi: 10.1136/bmj.k3788.

Yao C, Chen G, Song C, Keefe J, Mendelson M, Huan T, et al. Genome‐wide mapping of plasma protein QTLs identifies putatively causal genes and pathways for cardiovascular disease. Naturez Communications. 2018;9(1):3268. doi: 10.1038/s41467-018-05512-x.

Zanetti D, Tikkanen E, Gustafsson S, Priest JR, Burgess S, Ingelsson E. Birthweight, Type 2 Diabetes Mellitus, and Cardiovascular Disease: Addressing the Barker Hypothesis With Mendelian Randomization. Circulation: Genomic and Precision Medicine [Internet]. 2018;11(6). Available from: <https://www.ahajournals.org/doi/10.1161/CIRCGEN.117.002054>

**Results of the non-MR-Base systematic review.**

Studies included.

The search resulted in a total of 404 items. Of these, a random sample of 39 were reviewed. 4 were excluded because of the original exclusion criteria, a further 11 were excluded for being 2SMR studies which used MR-Base. This resulted in a random sample of 24, of which 15 were 2SMR and 9 were 1SMR studies.

Results.

The information from the studies included in this additional review can be found in **Supplementary Table 8**, in **Supplementary Table 9,** and **Figure 2.**

**Supplementary Table 10:** Studies included in additional review of non-MR-Base MR studies.

Bedard, A., S. J. Lewis, S. Burgess, A. J. Henderson and S. O. Shaheen (2018). Maternal iron status during pregnancy and respiratory and atopic outcomes in the offspring: a Mendelian randomisation study. BMJ Open Respir Res 5(1): e000275. doi: 10.1136/bmjresp-2018-000275.

Bergholdt, H. K. M., B. G. Nordestgaard, A. Varbo and C. Ellervik (2018). Lactase persistence, milk intake, and mortality in the Danish general population: a Mendelian randomization study. Eur J Epidemiol 33(2): 171-181. doi: 10.1007/s10654-017-0328-x.

Blauw, L. L., R. Li-Gao, R. Noordam, R. de Mutsert, S. Trompet, J. F. P. Berbee, Y. Wang, J. B. van Klinken, T. Christen, D. van Heemst, D. O. Mook-Kanamori, F. R. Rosendaal, J. W. Jukema, P. C. N. Rensen and K. Willems van Dijk (2018). CETP (Cholesteryl Ester Transfer Protein) Concentration: A Genome-Wide Association Study Followed by Mendelian Randomization on Coronary Artery Disease. Circ Genom Precis Med 11(5): e002034. <https://doi.org/10.1161/CIRCGEN.117.002034>

Caramaschi, D., A. E. Taylor, R. C. Richmond, K. A. Havdahl, J. Golding, C. L. Relton, M. R. Munafo, G. Davey Smith and D. Rai (2018). Maternal smoking during pregnancy and autism: using causal inference methods in a birth cohort study. Transl Psychiatry 8(1): 262. doi: 10.1038/s41398-018-0313-5.

Cheng, L., H. Zhuang, S. Yang, H. Jiang, S. Wang and J. Zhang (2018). Exposing the Causal Effect of C-Reactive Protein on the Risk of Type 2 Diabetes Mellitus: A Mendelian Randomization Study. Front Genet 9(NA): 657. doi: 10.3389/fgene.2018.00657.

Disney-Hogg, L., A. Sud, P. J. Law, A. J. Cornish, B. Kinnersley, Q. T. Ostrom, K. Labreche, J. E. Eckel-Passow, G. N. Armstrong, E. B. Claus, D. Il'yasova, J. Schildkraut, J. S. Barnholtz-Sloan, S. H. Olson, J. L. Bernstein, R. K. Lai, A. J. Swerdlow, M. Simon, P. Hoffmann, M. M. Nothen, K. H. Jockel, S. Chanock, P. Rajaraman, C. Johansen, R. B. Jenkins, B. S. Melin, M. R. Wrensch, M. Sanson, M. L. Bondy and R. S. Houlston (2018). Influence of obesity-related risk factors in the aetiology of glioma. Br J Cancer 118(7): 1020-1027.  doi: 10.1038/s41416-018-0009-x.

Estrada, K., C. W. Whelan, F. Zhao, P. Bronson, R. E. Handsaker, C. Sun, J. P. Carulli, T. Harris, R. M. Ransohoff, S. A. McCarroll, A. G. Day-Williams, B. M. Greenberg and D. G. MacArthur (2018). A whole-genome sequence study identifies genetic risk factors for neuromyelitis optica. Nat Commun 9(1): 1929. https://doi.org/10.1038/s41467-018-04332-3

Geng, T. T. and T. Huang (2018). Maternal central obesity and birth size: a Mendelian randomization analysis. Lipids Health Dis 17(1): 181. doi: 10.1186/s12944-018-0831-4.

Gill, D., M. K. Georgakis, M. Laffan, M. Sabater-Lleal, R. Malik, I. Tzoulaki, R. Veltkamp and A. Dehghan (2018). Genetically Determined FXI (Factor XI) Levels and Risk of Stroke. Stroke 49(11): 2761-2763. doi: 10.1161/STROKEAHA.118.022792.

Gomez-Acebo, I., T. Dierssen-Sotos, C. Palazuelos, P. Fernandez-Navarro, G. Castano-Vinyals, J. Alonso-Molero, C. Urtiaga, T. Fernandez-Villa, E. Ardanaz, M. Rivas-Del-Fresno, A. Molina-Barcelo, J. J. Jimenez-Moleon, L. Garcia-Martinez, P. Amiano, P. Rodriguez-Cundin, V. Moreno, B. Perez-Gomez, N. Aragones, M. Kogevinas, M. Pollan and J. Llorca (2018). Pigmentation phototype and prostate and breast cancer in a select Spanish population-A Mendelian randomization analysis in the MCC-Spain study. PLoS One 13(8): e0201750. oi: 10.1371/journal.pone.0201750.

Gottsater, M., G. Hindy, M. Orho-Melander, P. M. Nilsson and O. Melander (2018). A genetic risk score for fasting plasma glucose is independently associated with arterial stiffness: a Mendelian randomization study. J Hypertens 36(4): 809-814. doi: 10.1097/HJH.0000000000001646.

Harrison, S. C., M. V. Holmes, S. Burgess, F. W. Asselbergs, G. T. Jones, A. F. Baas, F. N. van 't Hof, P. I. W. de Bakker, J. D. Blankensteijn, J. T. Powell, A. Saratzis, G. J. de Borst, D. I. Swerdlow, Y. van der Graaf, A. M. van Rij, D. J. Carey, J. R. Elmore, G. Tromp, H. Kuivaniemi, R. D. Sayers, N. J. Samani, M. J. Bown and S. E. Humphries (2018). Genetic Association of Lipids and Lipid Drug Targets With Abdominal Aortic Aneurysm: A Meta-analysis. JAMA Cardiol 3(1): 26-33. doi: 10.1001/jamacardio.2017.4293.

He, L., I. Culminskaya, Y. Loika, K. G. Arbeev, O. Bagley, M. Duan, A. I. Yashin and A. M. Kulminski (2018). Causal effects of cardiovascular risk factors on onset of major age-related diseases: A time-to-event Mendelian randomization study. Exp Gerontol 107(NA): 74-86. doi: 10.1016/j.exger.2017.09.019. E

Isom, C. A., M. J. Shrubsole, Q. Cai, W. E. Smalley, R. M. Ness, W. Zheng and H. J. Murff (2019). Arachidonic acid and colorectal adenoma risk: a Mendelian randomization study. Clin Epidemiol 11(NA): 17-22. doi: 10.2147/CLEP.S186883.

Kia, D. A., A. J. Noyce, J. White, D. Speed, A. Nicolas, I. collaborators, S. Burgess, D. A. Lawlor, G. Davey Smith, A. Singleton, M. A. Nalls, R. Sofat and N. W. Wood (2018). Mendelian randomization study shows no causal relationship between circulating urate levels and Parkinson's disease. Ann Neurol 84(2): 191-199. doi: 10.1002/ana.25294.

Kunutsor, S. K., J. A. Laukkanen and S. Burgess (2018). Genetically elevated gamma-glutamyltransferase and Alzheimer's disease. Exp Gerontol 106(NA): 61-66. doi: 10.1016/j.exger.2018.03.001.

Larsson, S. C., S. Burgess and K. Michaelsson (2018). Genetic association between adiposity and gout: a Mendelian randomization study. Rheumatology (Oxford) 57(12): 2145-2148. doi: 10.1093/rheumatology/key229.

Meng, X. H., X. D. Chen, J. Greenbaum, Q. Zeng, S. L. You, H. M. Xiao, L. J. Tan and H. W. Deng (2018). Integration of summary data from GWAS and eQTL studies identified novel causal BMD genes with functional predictions. Bone 113(NA): 41-48. doi: 10.1016/j.bone.2018.05.012.

Nagel, M., P. R. Jansen, S. Stringer, K. Watanabe, C. A. de Leeuw, J. Bryois, J. E. Savage, A. R. Hammerschlag, N. G. Skene, A. B. Munoz-Manchado, T. andMe Research, T. White, H. Tiemeier, S. Linnarsson, J. Hjerling-Leffler, T. J. C. Polderman, P. F. Sullivan, S. van der Sluis and D. Posthuma (2018). Meta-analysis of genome-wide association studies for neuroticism in 449,484 individuals identifies novel genetic loci and pathways. Nat Genet 50(7): 920-927. doi: 10.1038/s41588-018-0151-7.

Ong, J. S., D. L. Hwang, V. W. Zhong, J. An, P. Gharahkhani, P. A. S. Breslin, M. J. Wright, D. A. Lawlor, J. Whitfield, S. MacGregor, N. G. Martin and M. C. Cornelis (2018). Understanding the role of bitter taste perception in coffee, tea and alcohol consumption through Mendelian randomization. Sci Rep 8(1): 16414. https://doi.org/10.1038/s41598-018-34713-z

Schooling, C. M., S. Luo and G. Johnson (2018). ADAMTS-13 activity and ischemic heart disease: a Mendelian randomization study. J Thromb Haemost 16(11): 2270-2275. doi: 10.1111/jth.14267.

Wang, N., J. Cheng, Z. Ning, Y. Chen, B. Han, Q. Li, C. Chen, L. Zhao, F. Xia, D. Lin, L. Guo and Y. Lu (2018). Type 2 Diabetes and Adiposity Induce Different Lipid Profile Disorders: A Mendelian Randomization Analysis. J Clin Endocrinol Metab 103(5): 2016-2025. doi: 10.1210/jc.2017-02789.

Yang, Q., S. L. Lin, M. K. Kwok, G. M. Leung and C. M. Schooling (2018). The Roles of 27 Genera of Human Gut Microbiota in Ischemic Heart Disease, Type 2 Diabetes Mellitus, and Their Risk Factors: A Mendelian Randomization Study. Am J Epidemiol 187(9): 1916-1922. doi: 10.1093/aje/kwy096.

Teumer, A., G. Gambaro, T. Corre, M. Bochud, P. Vollenweider, I. Guessous, M. E. Kleber, G. E. Delgado, S. Pilz, W. Marz, C. L. K. Barnes, P. K. Joshi, J. F. Wilson, M. H. de Borst, G. Navis, P. van der Harst, H. J. L. Heerspink, G. Homuth, K. Endlich, M. Nauck, A. Kottgen, C. Pattaro and P. M. Ferraro (2018). Negative effect of vitamin D on kidney function: a Mendelian randomization study. Nephrol Dial Transplant 33(12): 2139-2145. doi: 10.1093/ndt/gfy074.

**Supplementary Table 11:** number of studies indexed in PubMed when the search term was implemented on 01/06/2021

| Year | Count |
| --- | --- |
| 2021 | 246 |
| 2020 | 288 |
| 2019 | 155 |
| 2018 | 77 |
| 2017 | 49 |
| 2016 | 30 |
