## Supplementary Methods for "Investigating the transparency of reporting in two-sample summary data Mendelian randomization studies"

### **Search strategy**

The search terms for the bibliographic databases were as follows:

((“mr-base”) OR (“mr base”) OR (“mrbase”) OR (“TwoSampleMR”) OR (“Two Sample MR”) OR (“2SMR”) OR (“two sample mendelian randomization”) OR (“2 Sample MR”) OR (“two sample mendelian randomisation”) OR (“2 sample mendelian randomization”) OR (“2 sample mendelian randomisation”) OR (“Two-Sample MR”) OR  (“two-sample mendelian randomization”) OR (“2-Sample MR”) OR (“two-sample mendelian randomisation”) OR (“2-sample mendelian randomization”) OR (“2-sample mendelian randomisation”))

Here, ‘OR’ stands for the correct representation of a Boolean ‘or’ operator and each term encoded in the quotation marks is searched as a keyword.

**Modifications to the protocol**

Rather than having all disagreements in study selection arbitrated by a third researcher, initial disagreements were first discussed between the BW and NC who conducted the extraction. Likewise, instead of JY and RR arbitrating all disagreements in data-extraction, disagreements were instead resolved by the extractors re-reading the paper and coming to a joint decision.

Because coming to a decision over if an item had been reported with sufficient detail, or reported with insufficient detail, was decided to be too subjective, items were graded using a binary score: reported/not-reported.

We decided not to explore the either “The prevalence of reporting in each domain of the checklist” or “the proportion of studies which reported 100% and >50% of items in each individual domain, and in all domains”, from the original analysis plan, prior to running any analysis primarily because it was felt that, on reflection, these additional outcomes would not communicate significantly more information than what was already available from the other outcomes, but that providing an overall summary statistics (the mean) would improve the ability to summarise the overall level of reporting. The Supplementary Analyses were added after consultation with GDS, GH, MM, and JH.

**Description of review of non-MR-Base studies (as part of STROBE-MR development)**

Eligibility criteria

Studies were included if they were applied research articles using MR, so methodology articles, books, reviews, animal studies, brief reports, commentaries, editorials/corrections and letters to editors were excluded. For the purpose of this study, we additionally excluded 2SMR studies that used MR-Base to avoid overlapping studies.

Information sources and search terms

The search term: “MR[tiab] & 2018[PDAT]” was implemented in PubMed on the 1^st^ of April 2019.

Study selection

Eligibility was originally assessed by VS, with BW excluding MR-Base paper.

Data extraction

Each study had information extracted by two authors independently (from among VS, RR, BW, and JY). Conflicts were resolved through discussion between the extracting authors.

Reporting checklist

Items in the original extraction form was taken from the items in the original STROBE checklist, with sub sections added for MR specific factors (Supplementary Table 2). Of these, 11 had sufficient information for them to be harmonised with the questions extracted for this study. These included: both questions from the harmonisation domain, both questions from the sample overlap domain, the three questions on the core IV assumptions, the item on independence of SNPs, the criteria used to select SNPs, the primary model used, and the question on directionality.

Supplementary Table 1: Glossary of Mendelian Randomisation technical terms

| **Term** | **Definition** |
| --- | --- |
| Phenotype | Trait of interest in a genetic study. |
| GWAS | A genome wide association study (GWAS) is a hypothesis free genetic study design in which the phenotype is regressed on each genetic variant which has been measured. These typically have stringent corrections for multiple testing. |
| Genetic variant | A section of a DNA sequence which can vary within a population. In 2SMR setting this is generally a SNP (single nucleotide polymorphism). |
| SNP or single nucleotide polymorphism | When the genetic variant is a single base pair in the DNA in population. The possible base pairs are called alleles. SNPs are typically defined to exclude situations where the less common allele has prevalence less than 1%. |
| Harmonisation | By definition a SNP has more than one possible allele. If the two studies select different alleles as the effect allele, then the direction of association in the MR analyse will be incorrect.  Harmonization is the process of formatting the GWAS statistics to ensure that both studies use the same allele as the effect allele. |
| Palindromic SNPs | An individual’s genotype is made of two double helixes, one form each chromosome. When recording data, one side of each genetic data is read as the forward strand, and the other is read as the reverse strand (resulting in two forward and two reverse strands). A palindromic SNP is a SNP in which the same two alleles are found on both forward strands, and both reverse strands (e.g. C/G on the forward and G/C on the reverse). This can cause problems in harmonisation, because it is difficult to establish which is the minor allele in cases were both alleles have similar frequency. |
| GWAS QC (or quality control) | The, often standardised, process of removing potentially erroneous or biasing observations (individuals or SNPs) from a GWAS analysis. |
| Instrumental variable analysis | The statistical analysis used in a 2SMR design. The IV estimate in a 2SMR study uses the ratio of effect between a randomly assigned cause of the exposure on the outcome, over the effect of the random cause on the exposure. If the three assumptions below are valid, then this produces an unbiased estimate of the cause effect. |
| Instrument | The randomly assigned, or unconfounded, cause of the exposure used in IV analyses. |
| IV1: relevance | This assumption states that the instrument is associated with the exposure of interest. Bias in IV analyses is inversely proportional to the strength of association, measured using the F statistic. Instruments with an association of 10 or less are generally described as “weak instruments”. |
| IV2: independence | This assumptions states that the instrument-outcome association is not confounded. In an MR analysis, violations can occur if causes of non-random distributions of genetic variants also associate with the outcome of interest. Examples include assortative mating, dynastic effects, and population structure (described below). |
| IV3: exclusion restriction | This assumptions states that the exposure completely mediates the association between the instrument and the outcome. This assumption is often violated with genetic instruments due to pleitropy (variants being causally associated with many traits). 2SMR analyses will typically include various sensitivity analyses which can be valid when the exclusion restriction assumption is violated, but make other, weaker, assumptions instead. |
| IV4: Homogeneity/monotonicity/ constant effect | As with a randomised controlled trial, a point estimation in an MR analysis requires assuming that the effect estimate is homogeneous (the same in all participants) or monotonic (goes in the same direction in all participants). |
| Gene-environment equivalence | The assumption in MR that the effect of changes to the genotype on a phenotype can be analogous to changes of the environment on the phenotype (also called genocopy or phenocopy). |
| LD or linkage disequilibrium | Genetic variants are not completely independent of each other. Instead, each parental gamete will inherit random (haplotype) bocks of grandparental DNA. Therefore, the closer two variants are on the genome, the more likely they are to be inherited together. This causes a correlation between variants, called linkage disequilibrium. Because methods of combining SNPs (e.g. inverse variance waiting) assume that each SNP is independent of each other, the non-independence of SNPs had to be adjusted for or non-independent SNPs removed (see Clumping and Pruning below). |
| Proxy variant | GWASs often do not measure all variants, but only a sample. The existence of variants in strong linkage disequilibrium (i.e. highly correlated) means that proxies (proxy variants) of the required variant may be available if the two studies used in a 2SMR do not measure the same variants. Decisions about which variants can be used as proxies requires data from the relevant population with the entire genome, called a reference panel. |
| Directionality in MR | Germline genetic material is fixed at birth, and therefore reverse causation of genetic effects is impossible. However, MR studies can still suffer from directionality issues when the outcome is more proximal than the exposure, or if there is a bi-directional effect between the exposure and the outcome. |
| Genetic liability to the exposure | Binary traits with many genetic variants associated with them have an underlying continuous genetic liability which is measured in a GWAS. Because an individual with a high genetic liability to the trait may not develop it, MR studies with binary exposures are better described as using the genetic liability for the binary trait as the exposure. |
| GWAS weights | The effect estimates derived from a GWAS |
| Discovery sample | The sample in which SNP-phenotype associations are discovered. |
| Replication sample | The sample, ideally distinct from the discovery sample, in which the SNPs effect estimates for the IV analysis are estimated. Separate samples are often used in order to reduce over fit. |
| Colocalization | Genetic loci can associate with more than one trait. Colocalization is the process of attempting to determine if this is due to two SNPs at the same loci independently causing the two traits, or a pleiotropic effect of a single SNP on both traits. |
| Pleiotropy | Pleiotropy occurs when a genetic variant is causally associated with more than one trait. There are two types of pleiotropy: horizontal pleiotropy occurs when the two traits are independently caused by the variant, and vertical pleiotropy occurs when the causal association between the two traits occurs on the same path (e.g. trait one is a cause or cause of trait two) |
| Dynastic effects | When the parental genotype influences the offspring, typically independent of the offspring’s genotype. E.g. when the non-inherited part of the parental genotype has an effect on the environment which also impacts on the offspring (also called “passive gene-environment interaction”, and “genetic nurture” ). |
| Assortative mating | When individuals choose partners based on phenotypic manifestations of heritable traits, creating a correlation in their genotype. It can either be single-trait, e.g. if taller women partner with taller men, or cross-trait, e.g. if more educated men partner with taller women. Cross-trait assortment on the exposure and outcome, and some cases of single-trait assortment, will bias MR estimates. |
| Canalisation | When a phenotype remains invariant to change in a genetic variant associated with it, often due to the development of compensatory developmental processes. |
| Population stratification | The non-random allocation of genetic variants due to different localities, or sub populations, having different ancestries. This can cause genetic variants with a higher frequency in a subpopulation to correlate with a trait also more common in that population in the absence of a causal association. |
| Principle components of ancestry | In order to control for population stratification, many studies will control for the first e.g. ten components of principal component analyses generated from the genetic inter-relatedness of the sample. |
| BOLT-LMM | A type of linear mixed model used in genetic analyses as a way of controlling for population stratification. |
| Incident or prevalent cases | A case is prevalent if they developed the outcome either prior to recruitment into the study, or during study follow up. A case is incident if they entered the study without the outcome, and developed it during the study. Because prevalent cases require both developing the outcome, and surviving to study recruitment with it, factors which influence survival with the outcome can cause bias. |
| Clumping and pruning | Methods for removing SNPs which are correlated with each other (and therefore not independent) due to linkage disequilibrium. |

| **Item** | | | **Description** |
| --- | --- | --- | --- |
| 1 Tit & Abs | | | Do they state that it is an MR study in the title/abstract |
| 2 Background | | | Do they provide a background |
| 3 Objectives | | | Do they state the objectives, with a clearly causal hypothesis and direction of effect |
| 4 Design | **1SMR** | **2SMR** | Do they specify which type of MR study was conducted? |
|  | a) setting | a) studies | 1SMR: do they describe the setting of the study, 2SMR, do they describe the studies from which summary data was taken |
|  | b) participants | b) same population | 1SMR: do they provide a description of the participants (e.g. inclusion criteria), 2SMR: do they provide information on participants are drawn from the same sample) |
|  | c) data | c) samples | 1SMR: do they explain how data was collected, 2SMR: do they provide information on sample overlap |
|  |  | d) adjustment | What adjustments were made in the analysis? |
| 5 Variables | a) X, Y, Z | | Do they define the relevant variables? |
|  | b) Genetic instrument | | How were the genetic instruments and beta weights chosen |
| 6) Data Quality | a) imputation, low MAF | | Do they provide information on any imputation / MAF or HWE QC |
|  | b) palindromes | | How were palindromes dealt with? |
|  |  | c) harmonization | 2SMR: How was the data harmonised? |
| 7 Bias | | | Do they mention bias, and if so, which ones? |
| 8 Stat. Methods | a) target / statistics | | What is the estimator (e.g. two stage least square regression, Wald ratio etc) |
|  | b) independence | | Were the SNPs independent of each other |
|  | c) check MR assumptions | | Which of the three core IV assumptions were reported/well reported? |
|  | d) address violations | | How were violations of the core assumptions addressed |
|  | e) other sensitivity analyse | | Describe which sensitivity analysis were preformed |
|  | f) plots | | Describe plots presented? |
|  | g) reverse causation | | Describe what was done to address the possibility of reverse causation (e.g. bidirectional or Stilgier filtering) |
|  | h) additional analyses | | How were any additional analyses presented? |
|  | i) missing data |  | What was done to address any missing data issues? |
| 9 Descriptive | a) participants | a) GWAS | 1SMR: do they describe the participants were sampled?, 2SMR: do they describe the GWAS’s sampling procedure? |
|  | b) descriptive statistics | | Do they provide any descriptive statistics? |
|  | c) missing | | Do they describe any missing data |
|  | d) outcome | | Do they provide information on the outcome? |
| 10 Main Results | a) SNPs | | Do they provide an F statistic for the association between the genetic variant and the exposure |
|  | b) X-Y | | Do they report the beta values for the I-X or I-Y associations |
|  | c) Assess. of MR assumptions | | what core IV assumptions were assessed? |
|  | d) Sensitivity analysis - MR assumptions | | What sensitivity analyses for the core IV assumptions were presented in the results? |
|  | e) plots | | What plots were presented in the results? |
|  | f) other sensitivity | | What other sensitivity analysis were presented in the results? |
|  | g) reverse causality | | How was reverse causality explored in the results? |
|  | h) additional analyses | | How did they address any additional assumptions in the results? |
|  | i) translation | | Do they provide any information which would be helpful for translational research? e.g. units for the effect estimate. |
| 11 Results | | | Do they provide a summary of the main results |
| 12 Limit & strengths | | | Do they define the strengths and limitations? |
| 13 Interpretation | a) triangulation/ RCT? | | Do they attempt to triangulate or compare the results to an RCT? |
|  | b) mechanism | | Do they describe a mechanism |
|  | c) clinical relevance | | Do they explain if the results are clinically relevant? |
| 14 General. | | | Do they explain if the results will generalise? |
| 15 Future | | | What are the future directions? |
| 17 Data | | | Do they provide links to data sources? |
| Each is scored as 0 (not/poorly reported), 1 (partially reported), or 2 (adequately reported) | | | |

Supplementary Table 2: Items data was extracted on for the MR-STROBE review.
